# Pediatric Kidney Injury in Communities Impacted by Chronic Kidney Disease of Unknown Etiology (CKDu): A Comprehensive Systematic Review of Epidemiologic Studies

**DOI:** 10.1101/2024.09.12.24313469

**Authors:** Anna Strasma, Anisha Gerber, Isabela Agi Maluli, Elizabeth R. Blackwood, Sameera Gunasekara, P. Mangala C S De Silva, Nivedita Kamath, Marvin Gonzalez-Quiroz, Alison P. Sanders, Christina Wyatt, Nishad Jayasundara

## Abstract

**Background:** Chronic kidney disease of unknown etiology (CKDu) is a tubulointerstitial disease that disproportionately affects young, primarily male, agricultural workers in Mesoamerica and South Asia who lack traditional risk factors for kidney disease. Extensive research in adult populations suggests the etiology is complex and indicates that early childhood exposures could have an integral role.

**Objectives:** To systematically identify, summarize, and compare research in children living in CKDu endemic areas or with relevant CKDu-related exposures.

**Methods:** A systematic literature search was conducted in six databases for studies that report kidney health outcomes of pediatric populations living in proximity to CKDu-affected areas with no limitations on geography or study design. Studies were independently screened for inclusion and underwent quality assessment using the Appraisal Tool for Cross-Sectional Studies (AXIS) or the JBI Critical Appraisal Tool for Cohort Studies based on the study design by at least two authors. Data are compared narratively and graphically.

**Results:** We included twenty peer-reviewed publications and two meeting abstracts from eight different countries. The most common study design involved cross-sectional analysis of biological specimens from children in an established CKDu endemic area. Marked decreases in estimated glomerular filtration rate were generally not identified and prevalence of albuminuria differed widely between countries. Novel urinary biomarkers frequently demonstrated subclinical kidney damage, although the specific biomarker(s) varied between studies. Epidemiologic factors associated with evidence of subclinical kidney damage in children included proximity to agriculture or agrochemicals. Despite heterogenous study outcomes, all studies concluded that there were signs of kidney injury in children living in CKDu endemic areas or with a CKDu-relevant environmental exposure.

**Conclusion:** This systematic review suggests that the pathophysiologic process leading to CKDu may begin prior to adulthood. Future longitudinal research aimed at elucidating the multifaceted factors and exposures impacting entire communities, including children, is imperative for disease prevention strategies.

## Introduction

Geographically localized epidemics of chronic kidney disease of unknown etiology (CKDu) have emerged as a public health crisis over the past 30 years, causing significant morbidity and mortality in young and middle-aged adult agricultural laborers in Central America and Southeast Asia.^1,2^ While the underlying cause of chronic kidney disease (CKD) in these geographically disparate populations is unknown, the unifying feature is that it occurs primarily in agricultural communities.^1–3^

CKDu was first described in Sri Lanka, India, and Central America in the 1990s to 2000s.^4,5^ The high rates of mortality and morbidity associated with CKDu have been intensified by the silent progression of disease, lack of healthcare infrastructure, challenges in diagnosis of the disease at early stages, and lack of access to kidney replacement therapy (KRT) in socioeconomically disadvantaged populations.^4,6^ For example, in El Salvador kidney disease is the second leading cause of death in men, and in Sri Lanka’s North Central Province, there are 20,000 annual CKDu deaths.^1,7,8^ In Nicaragua, the community prevalence of CKD is reported as high as 42% in males and 10% in females.^9^ This disease often affects men of working age, who frequently provide the sole income for their families, and may be unable to work due to their disease. This has amplified the socioeconomic burden of CKDu leading to considerable community-wide economic consequences.

CKDu occurs in the absence of traditional risk factors that commonly cause kidney disease such as hypertension and diabetes.^1^ It is more prevalent among males, although females are also affected, and is typically diagnosed between 20-50 years old.^1,2^ Although kidney biopsies are not commonly performed in resource-limited regions, CKDu is characterized histologically as a chronic tubulointerstitial nephropathy with glomerulosclerosis.^10,11^ Classic features include progressive loss of glomerular filtration rate (GFR) with limited or no proteinuria and no hematuria.^1,2,12^ Hypokalemia and hypomagnesemia have also been described.^12,13^ Clinical symptoms are generally absent until late-stage disease, and there are no known treatments except KRT once patients progress to kidney failure.^2,12^

Despite two decades of research, the etiology of CKDu remains unclear. A complex and multifactorial causation is likely, involving environmental exposures to agrochemicals, both anthropogenic and geogenic metals, and recurrent heat stress and dehydration.^1,2,12,14^ Exposure to pesticides and other agrochemicals is known to induce acute kidney injury (AKI), although a specific causative agent for CKDu remains unidentified.^15,16^ While some metals are associated with CKD at high exposure levels, metal levels in drinking water and in human urine are highly variable in endemic regions, and usually exist within the recommended ranges or reference intervals respectively.^17,18^ Heat stress and dehydration have also been implicated given the location of the CKDu epidemics in tropical regions with high temperatures that are increasing further due to global warming.^19–23^ Environmental nephrotoxic exposures combined with unhealthy working conditions in agriculture may precipitate heat stroke and AKI – both of which may lead to CKD.^24^ Cumulative effect of metals, agrochemicals, and heat may exacerbate the disease.^18,25,26^ Genetic susceptibility might also contribute, as CKDu often clusters within families, but neither rare nor common variants have thus far been rigorously investigated.^14,27^ Underlying the multifactorial nature of the disease are important disparities in socioeconomic determinants of health within affected communities.^28^

Most CKDu literature focuses on agricultural or other manual labor workers with limited studies directly assessing environmental exposures and kidney outcomes in children.^13^ The assumption behind this approach is that children have healthy kidneys until they enter the workforce, where they encounter kidney insults that lead to CKDu. However, the identification of CKDu cases in adults in their early twenties implies that kidney damage may commence before employment.^29,30^ All the hypothesized exposures are prevalent throughout the community, and children may face higher risks from some exposures due to their increased surface area to body ratio and higher metabolic rate.^31^ Children are also undergoing critical periods of growth—including *in utero*, early childhood, and puberty—and exponential epigenetic maturation, which can be adversely impacted by environmental stress.^32–34^

Premature birth and low birth weight are known risk factors for CKD, and there is emerging evidence that environmental exposures *in utero* may affect fetal growth and kidney development.^35–37^ Many pesticides and metals cross the placenta and reach the fetus where they could have nephrotoxic effects.^38–40^ A mixture of metals and herbicides identified in Sri Lankan drinking water was found to affect kidney development in animal models.^41^ Collectively, altered trajectories during key developmental periods may lead to increased susceptibility to CKD later in life.^41,42^ Accordingly, there is a small but growing body of literature evaluating how environmental exposures implicated in CKDu may begin causing kidney damage prior to adulthood.

The potential for childhood kidney damage is substantiated by several endemic nephropathies that serve as historical corollaries to CKDu. For example, in the early 1900s, an epidemic of chronic tubulointerstitial nephritis among children and adults in Queensland, Australia, was linked to lead paint ingestion from outdoor veranda railings.^43,44^ Balkan endemic nephropathy, first described in the 1950s as a form of chronic tubulointerstitial nephritis, is caused by ingestion of plants containing aristolochic acid.^45^ While primarily described in adults, children from affected families exhibited higher tubular proteinuria levels compared to children in unaffected families.^46^ Historically, evaluation of childhood kidney health has been crucial for identification of relevant exposures and primary and secondary prevention strategies.

Conventional markers of kidney function (e.g., albuminuria, serum creatinine, estimated GFR) have limited sensitivity and specificity, particularly for detection of kidney injury and CKD in very early stages. A detectable rise in serum creatinine is a relatively late sign of CKD progression.^47^ While albuminuria is generally used as a screening tool for pediatric CKD, it is not an expected feature in early CKDu cases, and community albuminuria screening may underestimate CKDu prevalence.^12,48^

Several urinary biomarkers show promise in detecting kidney damage prior to detectable changes in serum creatinine or urinary albumin.^49^ In adults, kidney injury molecule 1 (KIM-1) and neutrophil gelatinase-associated lipocalin (NGAL), may hold potential for early diagnosis of kidney injury in CKDu endemic areas.^50^ Urinary biomarkers have potential for early detection of pediatric AKI and CKD, although most are not yet approved for clinical diagnosis.^49^ KIM-1, NGAL, N-acetyl-beta-glucosaminidase (NAG), and interleukin-18 (IL-18) are biomarkers indicative of tubulointerstitial injury while monocyte chemoattractant protein-1 (MCP-1) and chitinase-3-like protein-1 (YKL-40) are markers of inflammation.^49^ Urinary biomarkers may be sensitive and specific tools for identifying childhood kidney damage in CKDu-impacted communities, as discussed in several studies included in this review. In addition, advances in urinary proteomics, metabolomics and epigenomics hold promise for development of improved CKDu-diagnostic biomarkers or panels of biomarkers.^51–53^

The purpose of this systematic review is to identify, summarize, and compare the important research being conducted in children living in CKDu endemic areas or with relevant CKDu-related exposures. We aim to use this information to help guide further research, optimize screening, and advocate for children’s kidney health around the globe.

## Methods

### Protocol and Search Strategy

The protocol for this study was registered in the PROSPERO database for systematic reviews (ID CRD42023394987) where it can be accessed. The search was developed and conducted by a professional medical librarian (EB) in consultation with the rest of the authors and included a mix of keywords and subject headings representing chronic kidney disease of unknown etiology, Mesoamerican nephropathy, chronic interstitial nephritis of agricultural communities, chronic kidney disease, agriculture, farm, and pediatrics. No filters were used to limit the results by language or geographic area. The searches were independently peer reviewed by another librarian using a modified Peer Review of Electronic Search Strategies (PRESS) Checklist. Searches were performed in six databases: MEDLINE via PubMed, Embase via Elsevier, CINAHL Complete via Ebsco, Cochrane via Wiley, Web of Science via Clarivate, and SciELO via FAPSEP-BIREME. The search was conducted on January 13, 2023 and returned 1,518 unique citations. Complete reproducible search strategies for all databases are detailed in the Supplementary Materials. All citations were imported first into EndNote and then into Covidence, a systematic review screening software that also removes duplicates. Template data collection forms and the extracted data can be provided by contacting the authors.

### Study Eligibility Criteria

Studies were included if they investigated children (< 18 years old) living in proximity to an area where the studies’ authors reported that CKDu affects adults or the studies’ authors investigated an exposure they deemed relevant to CKDu. All studies must report on kidney health outcomes. Articles that included both children and adults were only included if data on children were specifically reported or provided on author request. We excluded studies if the adult kidney disease discussed was not consistent with the case definition of CKDu, which is a chronic interstitial kidney disease lacking traditional risk factors for CKD such as diabetes or hypertension. Only articles in English or Spanish were included. The search was not limited by geographic area or by study design.

### Study Selection

All titles and abstracts were independently evaluated for inclusion by two authors (AS and AG). Any disagreement was adjudicated by independent review by a third author (NJ). All full-text articles were assessed independently using the same criteria by two authors (AS and AG), and any disagreement on inclusion of the article was resolved by independent review of a third author (NJ). Inter-rater agreement was assessed using Cohen kappa coefficient.

### Data Extraction and Quality Assessment

A standardized data extraction form was used by AS and AG to extract study characteristics: study design, study location, location characteristics, study population characteristics, sample size, outcomes, conclusions, study strengths and limitations. Any difficulty in data extraction was discussed by AS, AG, and NJ with review of the original papers. AS and AG contacted authors directly for questions related to this review, which most often involved requesting data for a subgroup of pediatric participants. Each selected study was independently assessed by AS and AG using the Appraisal Tool for Cross-Sectional Studies (AXIS) or the JBI Critical Appraisal Tool for Cohort Studies based on the study design (supplemental material). The AXIS tool includes 20 total questions assessing the quality of the study divided into sections of Introduction, Methods, Results, Discussion, and Other. The JBI Critical Appraisal Tool for Cohort Studies includes 10 questions focused on the methods of the study. Neither tool includes a scoring system. Rather, the tools were used as a general guide to independently assign studies a quality rating of high, medium, or low. Any disagreement in quality rating was discussed by reviewing authors to reach consensus. When only an abstract was available, it was excluded from quality assessment.

Data were extracted independently by at least two authors. Data were represented and compared narratively and graphically. No statistical comparisons or meta-analysis were performed given heterogeneity of the original articles.

## Results

### Study Selection

Our initial literature search identified 1,518 possible studies (Figure 1). Through title and abstract screening, this was narrowed down to 193 articles and abstracts that underwent full text review. One article published after the initial literature search was identified by authors and included. A total of 20 articles and 2 abstracts met the criteria and were included in this systematic review and summarized in Table 1. Inter-rater reliability for inclusion was high with Cohen’s Kappa of 0.76 at the stage of title and abstract review and 0.72 for full text review.

**Figure 1:**
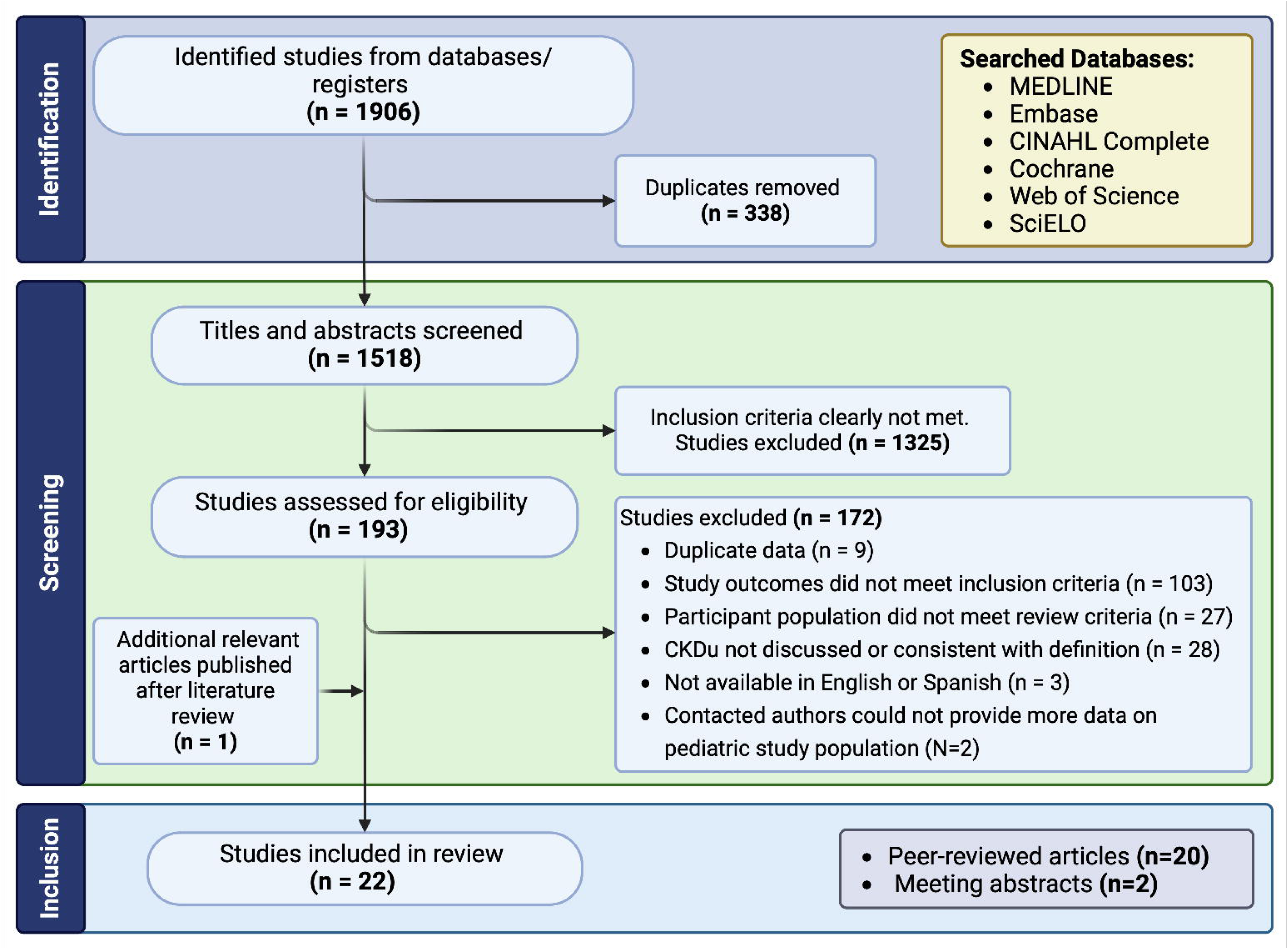
Flow diagram demonstrating inclusion and exclusion of studies for review.

**Table 1:** Characteristics, main results, and quality assessment (QA) scores of included studies. Study outcomes include the kidney-relevant outcomes reported in each study. Main relevant results were chosen and summarized using a standardized data extraction form. Quality assessment score is based on the Appraisal Tool for Cross-Sectional Studies (AXIS) or the JBI Critical Appraisal Tool for Cohort Studies depending on study design.

| Study Location and Title | First Author, Publication Year | Study Design | Sample size | Age Range (years) | Study Outcomes | Main Results | QA score |
| --- | --- | --- | --- | --- | --- | --- | --- |
| Sri Lanka (6) |  |  |  |  |  |  |  |
| *Early renal damage among children living in the region of highest burden of CKDu in Sri Lanka* | Agampodi<sup>56</sup>, 2018 | Cross-sectional | 2880 | 5-11 | UACR, serum creatinine, eGFR | 8.7% of children had albuminuria<br>Only 0.5% with UACR >300 mg/gCr.<br>Mean eGFR was significantly lower in the group with elevated UACR | Low |
| *The incidence, prevalence, and trends of CKD and CKDu in the North Central Province of Sri Lanka: an analysis of 30,566 patients* | Ranasinghe<sup>73</sup>, 2019 | Cohort | 30566 total, 564 children (<20 yo) | All ages | Prevalence, incidence, and mortality from CKD | CKD incidence increased over time in the total study population<br>CKD cases in the total study population are clustered around paddy fields and irrigation tanks and are not uniform across districts. | High |
| *Urinary biomarkers of renal injury KIM-1 and NGAL: Reference intervals for healthy pediatric population in Sri Lanka* | De Silva<sup>78</sup>, 2021 | Cross-sectional | 909 | 10-18 | UACR, uNGAL, uKIM-1 | Established reference intervals for uKIM-1 and uNGAL for girls and boys in Sri Lanka | Medium |
| *Urinary biomarkers indicate pediatric renal injury among rural farming communities in Sri Lanka* | Gunasekara<sup>66</sup>, 2022 | Cross-sectional | 804 | 10-18 | UACR, uNGAL, uKIM-1 | Participants in CKDu endemic regions had elevated urinary KIM-1 expression compared to other regions. Urinary NGAL did not show this regional variation.<br>Albuminuria was rare (1-2%) in all regions but was associated with higher urinary KIM-1 values. | High |
| *Urinary Cystatin C: pediatric reference intervals and comparative assessment as a biomarker of renal injury among children in the regions with high burden of CKDu in Sri Lanka* | Sandamini<sup>67</sup>, 2022 | Cross-sectional | 850 (phase 1), 892 (phase 2) | 10-17 | UACR, urine cystatin C | Established reference intervals for urinary cystatin C in Sri Lanka.<br>Urinary cystatin C levels were not elevated in CKDu endemic regions compared to non-endemic regions.<br>Urinary cystatin C was positively correlated with UACR in endemic regions. | Medium |
| *Environmental heat exposure and implications on renal health of pediatric communities in the dry climatic zone of Sri Lanka: An approach with urinary biomarkers* | Gunasekara<sup>77</sup>, 2023 | Cross-sectional | 697 | 12-16 | UACR, uNGAL, uKIM-1, effective temperature, wet-bulb globe temperature, | No elevation of uKIM-1 or UACR in high heat exposure (outdoor sports) versus low heat exposure groups. uNGAL was higher in the high heat exposure group only in the nonendemic regions. | Medium |
|  |  |  |  |  | heat index, humidex | Irrespective of the degree of heat exposure, children in endemic areas had higher uKIM-1. uNGAL was higher in endemic regions for girls only and UACR did not differ between regions. |  |
| Nicaragua (4) |  |  |  |  |  |  |  |
| *Urine biomarkers of kidney injury among adolescents in Nicaragua, a region affected by an epidemic of CKDu* | Ramírez-Rubio<sup>68</sup>, 2016 | Cross-sectional | 200 | 12-18 | UACR, uNAG, uNAGL, uIL-18 | Urinary NGAL and NAG were higher in regions with higher CKDu risk while IL-18 did not show this regional variation.<br>Proteinuria was rare throughout. Girls had higher levels of uNGAL and uNAG than boys. | Medium |
| *eGFR, CKD epidemiology in a Nicaraguan high risk area for Mesoamerican Nephropathy (ABSTRACT ONLY)* | Sidoti<sup>74</sup>, 2018 | Cross-sectional | 1202 total | 12-22 | Serum creatinine and eGFR | Prevalence of decreased eGFR is high (0.75%)<br>Differences in eGFR were better discriminated with use of the full age spectrum equation for children compared to the Schwartz equation. | N/A |
| *Biomarkers of kidney injury among children in a high risk region for CKDu* | Leibler<sup>57</sup>, 2021 | Cross-sectional | 210 | 7-17 | Urine dipstick, uNGAL, uKIM-1, uIL-18, uMCP-1, uYKL-40. Serum creatinine. | 9% of children had eGFR <100 and 29% had eGFR >160.<br>Median urinary NGAL, IL-18, and KIM-1 concentrations exceeded the reference values from healthy children in other countries.<br>Urinary NGAL was associated with increasing age and dysuria. Girls had higher concentrations of all biomarkers than boys. | Medium |
| *Urinary findings among adults and children in a region of Nicaragua endemic for Mesoamerican Nephropathy* | Strasma<sup>58</sup>, 2022 | Cross-sectional | 471 total, 210 children | 0 – 17 | Urine dipstick and microscopy | Leukocyturia was common overall and was more common in adults than children.<br>Proteinuria and hemoglobinuria were rare. | Low |
| El Salvador (1) |  |  |  |  |  |  |  |
| *CKD in children and adolescents in Salvadoran farming communities: NefroSalva Pediatric Study (2009-2011)* | Orantes-Navarro<sup>59</sup>, 2016 | Cross-sectional | 2115 | 2-17 | Urine dipstick, UACR, serum creatinine. | Prevalence of urinary abnormalities was 4% largely based on microalbuminuria detection and did not differ between age groups or regions.<br>Glomerular hyperfiltration observed in all age groups and both sexes. | High |
| Guatemala (2) |  |  |  |  |  |  |  |
| *CKD among children in Guatemala* | Cerón<sup>70</sup>, 2014 | Cross-sectional | 1545 | <18 | Prevalence of CKD and ESKD, cause of disease (if known) | In national database of pediatric CKD patients, the cause of CKD was undetermined in 43%. These patients could have CKDu or another undiagnosed process leading to CKD.<br>CKD without a determined etiology had a higher<br>incidence in Guatemala City and Pacific Coast. | High |
| *Factors associated with CKD of non-traditional causes among children in Guatemala* | Cerón<sup>71</sup>, 2021 | Cross-sectional | 156 | <18 | Individual and household characteristics that could be risk factors for CKD without a determined etiology | 76% of pediatric patients with stage 5 CKD or ESKD in national database had CKD without a determined etiology. These patients could have CKDu or another undiagnosed process leading to CKD.<br>Compared to patients with CKD from known causes, risk factors for CKD without a determined etiology were lower level of maternal education and agricultural practices in their municipality of residence. | Medium |
| Mexico (6) |  |  |  |  |  |  |  |
| *Environmental exposure to arsenic and chromium in children is associated with KIM-1* | Cárdenas-González<sup>65</sup>, 2016 | Cross-sectional | 83 | 5-12 | Metal levels, uKIM-1, uNGAL, eGFR, UACR, miR-21 | Children had exceedingly high arsenic and chromium exposure based on analysis of urine, blood, and drinking water.<br>Higher urinary KIM-1 associated with higher chromium and arsenic while eGFR, NGAL, and miR-21 were not. These kidney injury biomarkers were not associated with cadmium, fluoride, or lead. | Medium |
| *Prevalence of albuminuria in children living in a rural agricultural and fishing subsistence community in Lake Chapala, Mexico* | Lozano-Kasten<sup>60</sup>, 2017 | Cross-sectional | 394 | <17 | UACR, serum creatinine, eGFR | The prevalence of albuminuria persistent across at least 12 weeks was markedly high at 46%.<br>In participants with persistent albuminuria, 78% had eGFR <60. | Medium |
| *CKD in children aged 6-15 years and associated risk factors in Apizaco, Tlaxcala, Mexico, a pilot study* | Ortega-Romero<sup>79</sup>, 2019 | Cross-sectional | 109 | 6-15 | Serum creatinine, UACR, renal ultrasound, biopsy (in 1 participant) | 5 participants (4.6%) had persistent urinary abnormalities for at least 6 months and were found to have stage 2 CKD based on eGFR.<br>Risk factors for CKD included male sex, older age, and living <1 kilometer from a river.<br>Renal ultrasound was normal in all CKD participants. One renal biopsy performed and revealed Alport syndrome. | High |
| *CKDu in Mexico: The case of Poncitlan, Jalisco* | Garcia-Garcia<sup>69</sup>, 2020 | Cross-sectional | 650 | 2-17 | Serum creatinine, urine dipstick, prevalence of KRT | Prevalence of proteinuria was 44% compared to 4% in other municipalities | Medium |
| *High prevalence of end-stage renal disease of unknown origin in Aguascalientes Mexico: role of the registry of CKD and renal biopsy in its approach and future directions* | Gutierrez-Peña<sup>72</sup>, 2021 | Cross-sectional | 2827 | All ages | Prevalence of ESKD, cause of ESKD, biopsy results | ESKD prevalence in this population of children is 33.3 per million population<br>For all renal biopsies performed in 1–10-year-olds, the most common histopathologic diagnoses were Minimal change disease, FSGS, and IgA nephritis. For 11-20yos, most common diagnosis was FSGS and Lupus. Tubulointerstitial nephritis was rare (<8% of biopsies in each age group). | High |
| *Histologic characterization and risk factors for persistent albuminuria in adolescents in a region of highly prevalent end stage renal failure of unknown origin* | Macias Diaz<sup>62</sup>, 2022 | Cross-sectional | 513 | 10-17 | Serum creatinine, eGFR, UACR, renal ultrasound, renal biopsy | Prevalence of persistent albuminuria over a median of 5 months was 3.7% and was associated with decreased total renal volume, living close to maize crops, use of pesticides at father's workplace, family history of CKD, and blood pressure abnormalities while breastfeeding and high BMI were protective.<br>Low kidney volume also associated with decreased eGFR.<br>18 kidney biopsies with common findings of glomerulomegaly, partial foot process effacement, microvillous degeneration.<br>Participants with persistent albuminuria were started on Losartan and had a significant decrease in their albuminuria. | High |
| Brazil (1) |  |  |  |  |  |  |  |
| *Environmental exposure and effects on health of children from a tobacco-producing region* | Nascimento<sup>63</sup>, 2017 | Cohort | 40 | 6-12 | UACR, uNAG, urine cotinine, measures of pesticide exposure | Noted higher albuminuria, urinary cotinine, chromium, and markers of oxidative damage in periods with higher pesticide exposure. | Medium |
| India (1) |  |  |  |  |  |  |  |
| *Prevalence of renal dysfunction in primitive tribal groups in Utnoor (Telangana) (ABSTRACT ONLY)* | Bukka<sup>75</sup>, 2021 | Cross-sectional | 1108, 651 children | All ages | Prevalence and etiologies of renal dysfunction | Prevalence of decreased eGFR among tribal groups in India is high with the maximum prevalence among subjects 6-10yo. | N/A |
| Egypt (1) |  |  |  |  |  |  |  |
| *Acute and cumulative effects of repeated exposure to chlorpyrifos on the liver and kidney function among Egyptian adolescents* | Ismail<sup>64</sup>, 2021 | Cohort | 119-279 depending on year | 12-21 | Serum creatinine, serum BUN, urine measurements of TCPy (marker for | Although the average serum creatinine was in normal range, they significantly increased immediately following exposure to chlorpyrifos in both in applicators and those residing in nearby communities. | Low |
|  |  |  |  |  | chlorpyrifos),<br>liver function<br>panel,<br>cholinesterase<br>activity | Serum urea levels did not recover to baseline after each chlorpyrifos application. |  |

Abbreviations: Quality Assessment (QA), Chronic kidney disease of unknown etiology (CKDu), urinary albumin to creatinine ratio (UACR), estimated glomerular filtration rate (eGFR), chronic kidney disease (CKD), years old (yo), kidney injury marker 1 (KIM-1), neutrophil gelatinase associated-lipocalin (NGAL), urinary (u), N-acetyl- $\beta$ -D-glucosaminidase (NAG), interleukin-18 (IL-18), monocyte chemoattractant protein-1 (MCP-1), and chitinase-like protein (YKL-40), end stage kidney disease (ESKD), kidney replacement therapy (KRT), focal segmental glomerulosclerosis (FSGS), body mass index (BMI)

### Assessment of Quality and Publication Bias

The quality assessment score for full-length studies is included in Table 1. Overall, 35% (7/20) were of high quality, 50% (10/20) were of medium quality, and 15% (3/20) were of low quality based on the appraisal tools utilized. The most common reasons studies were rated as medium or low quality were cross-sectional design, lack of guideline recommended diagnosis of CKD, or lack of robust statistical methods. Publication bias likely affects our result since it is possible that a manuscript describing a lack of kidney disease markers in children in CKDu-endemic areas may not be published.

### Study Populations and Characteristics

Most studies were performed in countries known to have a high burden of CKDu in adults (Figure 2A). A total of 13 studies were performed in Mesoamerica (1 in El Salvador, 4 in Nicaragua, 2 in Guatemala, 6 in Mexico) and 6 in Sri Lanka. One study each was identified in Brazil, Egypt, and India. Of note, there were 2 identified studies that included some pediatric participants, but they were not included in the review because no specific data was available for child participants when authors were contacted directly.^54,55^

**Figure 2:**
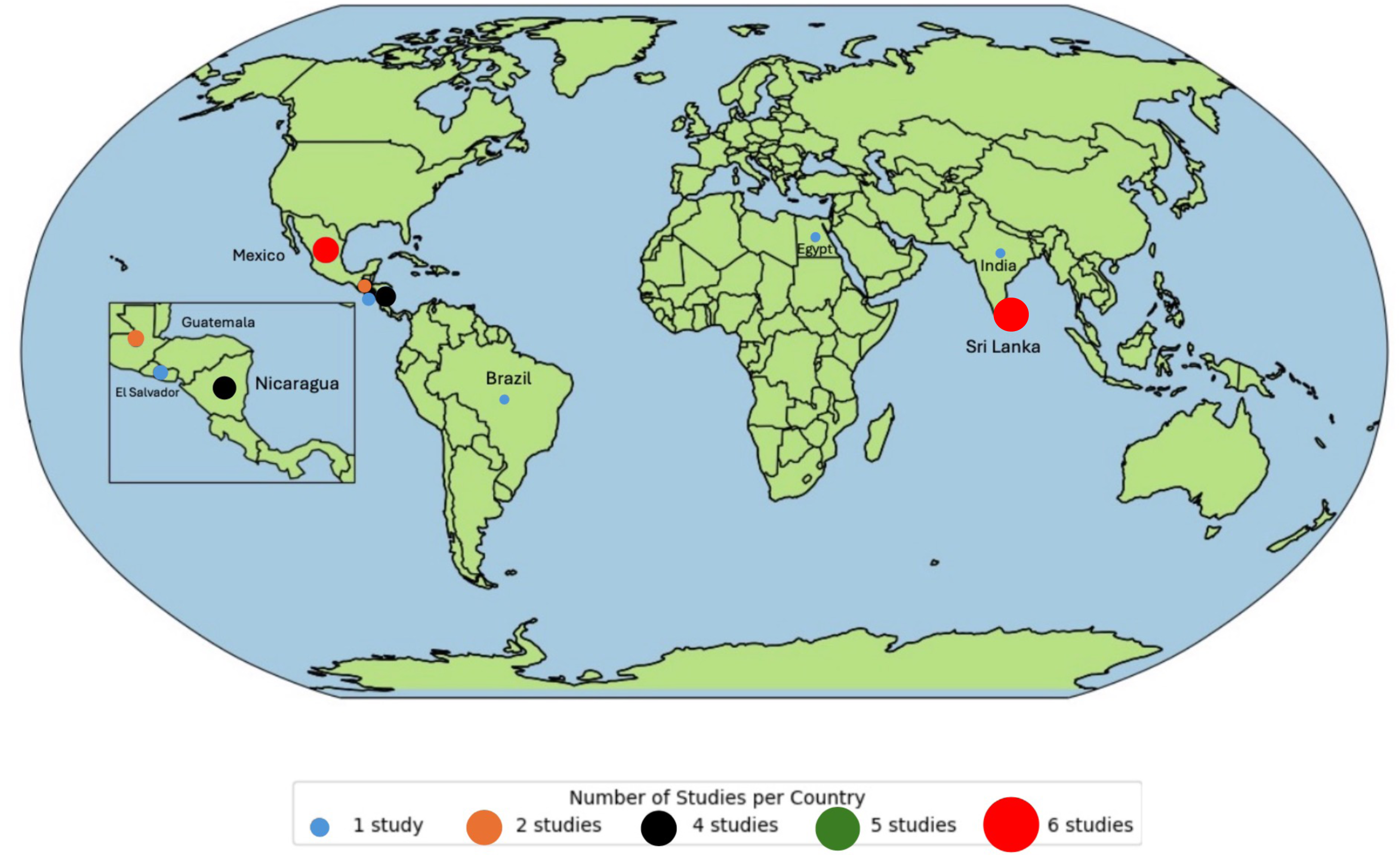
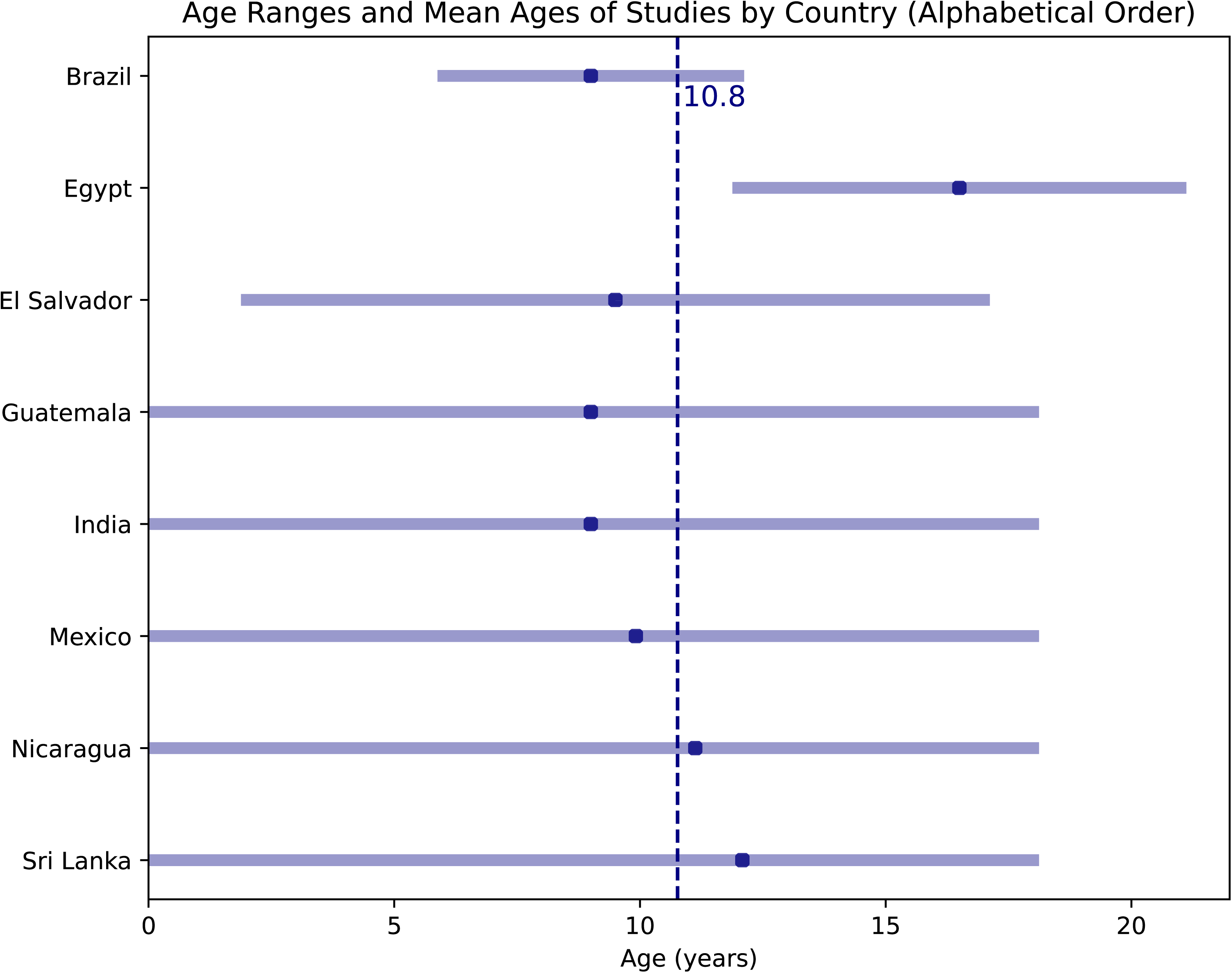
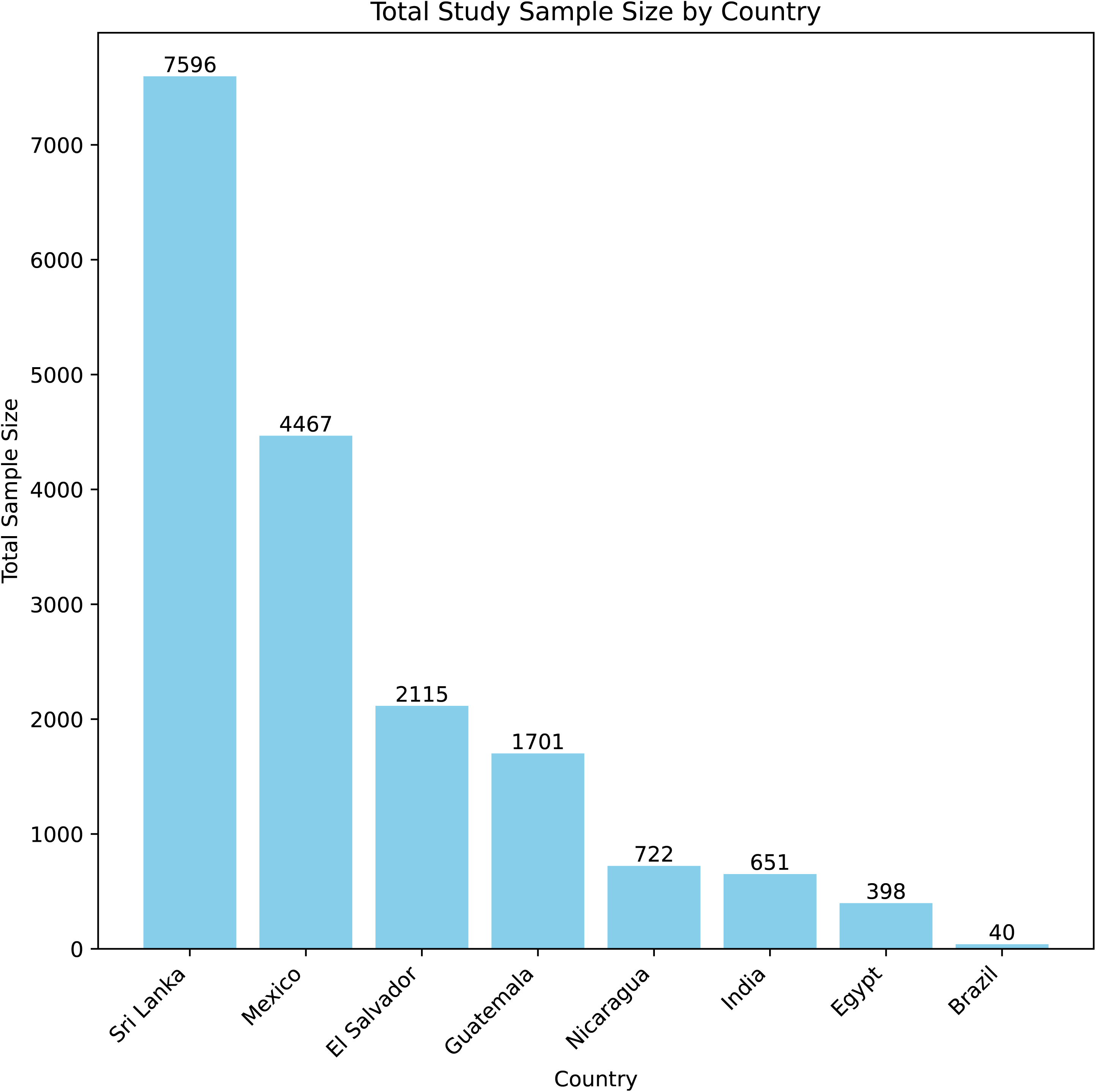
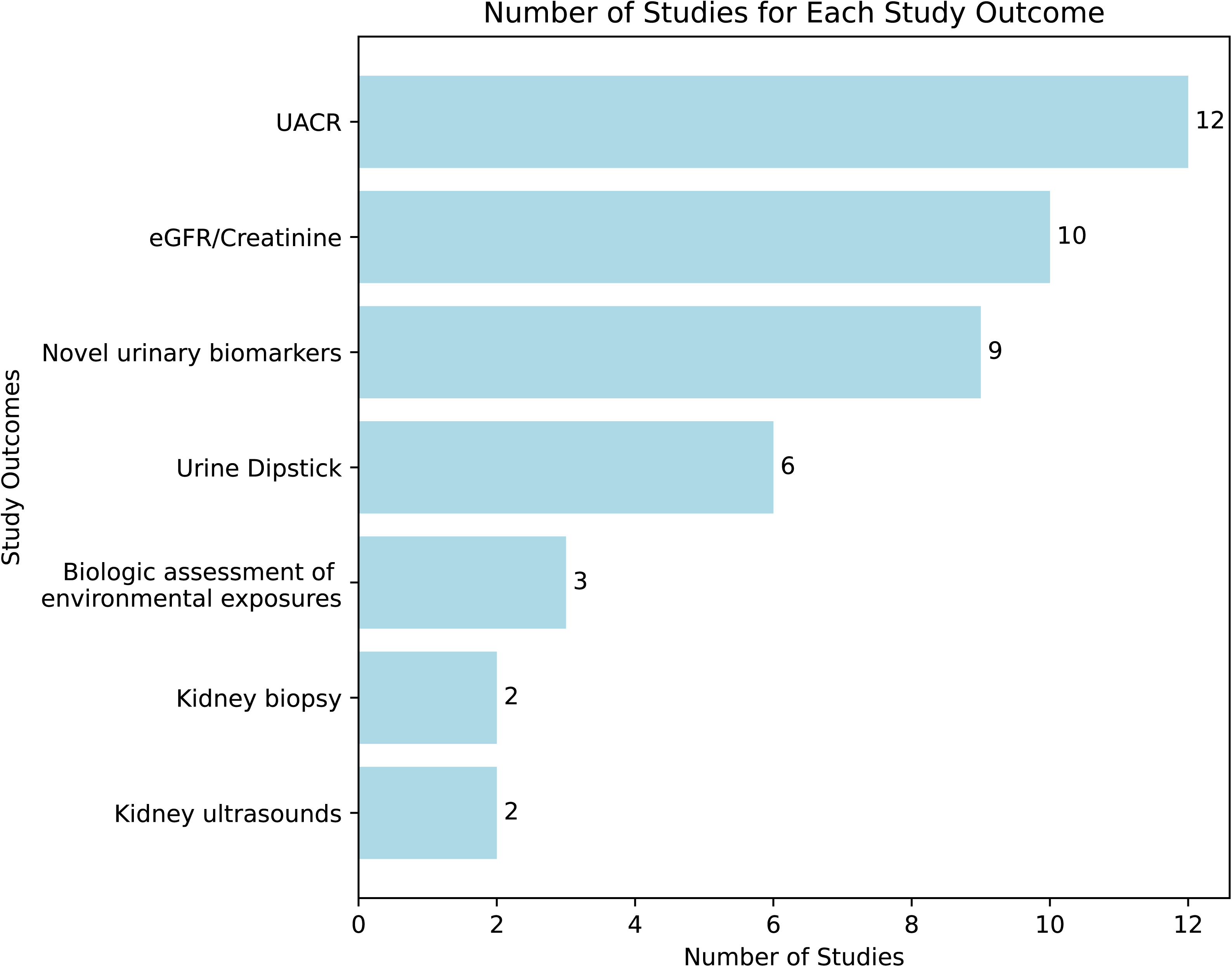
Characteristics of included studies (panel). A) Map and frequency of studies categorized by country. B) Mean age and full age range of children participants categorized by country. C) Sample sizes categorized by country. D) Frequency of study outcomes examined in included studies.

The most common study strategy involved collection of biological specimens from schools, communities, or the workplace in CKDu endemic area (16/20 studies). Kidney biomarker results were simply reported or were compared to reference values or literature review in many studies.^56–65^ Some studies compared results across regions with different levels of CKDu prevalence within the same country. ^66–69^

Seventeen studies were cross-sectional in design and 3 were cohort studies (Table 1). Cross-sectional studies included analyses of clinic or national registries for CKD or end stage kidney disease (ESKD) and analysis of risk factors in CKD clinic patients.^70–72^ There were two prospective cohort studies, which were both focused on environmental exposures that varied over time.^63,64^ Although the text of these two studies mentions CKDu, the studies’ objectives were to assess the impacts of possible nephrotoxic exposures in young people and were not necessarily focused on CKDu.^63,64^ Ranasinghe *et al.* conducted a retrospective cohort study following medical charts over time to calculate incidence and mortality from CKD.^73^

Twelve studies included school-age children as participants. Only 4 studies analyzed biological samples from children less than 5 years old.^58–60,69^ Some studies included both adults and children, and we focused this review on their findings in children.^58,72–75^ Ismail *et al.* included adolescents aged 12-21 years old who were occupationally exposed in both in the community and in the workforce. The mean age and range of ages included for each country are shown in Figure 2B. Participants were most frequently recruited from schools (12 studies) and less commonly recruited from community visits (6 studies), healthcare systems (2 studies), or the workplace (1 study). For studies that collected biological samples, the sample sizes tended to be larger in Sri Lanka (median 837 participants) compared to Mesoamerica (median 210 participants) (Figure 2C).

Of the 16 studies that collected biological samples, all of them analyzed urine and 9 analyzed serum (Figure 2D). Only 6 followed the Kidney Disease Improving Global Outcomes (KDIGO) CKD diagnosis guidelines and collected 2 samples at least 3 months apart.^59–64,76^ The most common urine analyses were urine albumin to creatinine ratio (UACR) (12 studies) and urine dipstick (6 studies). The most common serum analysis was serum creatinine, which was then used to calculate the estimated GFR (eGFR). In all studies reporting eGFR, it was calculated using the Schwartz bedside equation. One abstract by Sidoti *et al.* reported that the Full Age Spectrum eGFR Equation may be the preferred method in a CKDu endemic area of Nicaragua.^74^ Kidney ultrasounds and kidney biopsies were only performed in two studies.^61,62^ Environmental exposures to metals, agricultural products, or pesticides were measured in 3 studies.^63–65^ One study collected wet bulb globe temperatures to estimate the heat index as a proxy of outdoor heat exposure.^77^

Urinary biomarkers were studied in 8 studies, but the type of biomarkers tested varied between studies and countries. NGAL (6 studies) and KIM-1 (5 studies) were the most frequently studied urinary biomarkers. Other urinary biomarkers evaluated include NAG (2 studies), IL-18 (2 studies), cystatin C (1 study), MCP-1 (1 study), and YKL-40 (1 study). Biomarker results were not presented in a standardized manner across studies and varied based on sex and age. In addition, these tests generally have not been validated in the study population, with the exception of Sri Lanka where pediatric reference values for urinary(u) KIM-1, NGAL, and cystatin C were described by studies included in this review.^67,78^

### Heterogeneity in Measured Kidney Health Outcomes

In all studies, the authors concluded that children in CKDu endemic areas or with CKDu-relevant exposures may show signs of early kidney damage despite the heterogeneity in approaches and study designs. A summary of main results are shown in Table 1.

The prevalence of decreased eGFR differed between studies and endemic regions. In El Salvador, Orantes-Navarro *et al.* identified a low prevalence (0.3%) of decreased eGFR for at least 3 months.^59^ Leibler *et al.* in Nicaragua identified 4% of participants with decreased eGFR (<90 ml/min/1.73m^2^) on a single measurement.^57^ They also noted 29% of participants had eGFR measurements ≥ 160 ml/min/1.73m^2^, which could be a marker of glomerular hyperfiltration.^57^ In contrast, Agampodi *et al.* reported a markedly high prevalence of decreased eGFR in participants with albuminuria (55%) as well as a control group without albuminuria (39%) based on a single measurement in Sri Lanka.^56^ Additionally, Lozano-Kasten *et al.* observed that 99% of participants with persistent albuminuria had decreased eGFR.^60^ Cárdenas-González *et al.* identified no association between eGFR and levels of arsenic, chromium, or other environmental contaminants.^65^

Albuminuria results were inconsistent across studies and highly variable based on study country. Markedly high prevalence of elevated UACR was reported in multiple studies across central Mexico. More than 40% of studied children had albuminuria or proteinuria in the municipality of Poncitlan and in one rural community in Lake Chapala.^60,69^ Agampodi *et al.* identified a high prevalence of albuminuria in children in an endemic region of Sri Lanka and found that mean eGFR was lower in the group with albuminuria compared to the group without albuminuria.^56^ On the other hand, Gunasekara *et al.* (Sri Lanka), Sandamini *et al.* (Sri Lanka), and Ramírez-Rubio *et al.* (Nicaragua) did not identify increased UACR in high-risk regions compared to low-risk regions.^66–68,77^ Orantes-Navarro *et al.* (El Salvador), Leibler *et al.* (Nicaragua), and Strasma *et al.* (Nicaragua) reported low prevalence of albuminuria or proteinuria in endemic regions.^57–59^

Studies with novel urinary biomarkers reported mixed findings on the differential prevalence of sub-clinical kidney damage in communities with varying burden of CKDu. In two studies by Gunasekara *et al.* (2022, 2023) reported that uKIM-1 levels were higher in children with albuminuria and in children residing in CKDu high-risk regions compared to those in nonendemic regions in Sri Lanka.^66,77^ However, they did not observe regional variation of uNGAL^66^. Ramírez-Rubio *et al.* identified higher levels of uNGAL and uNAG in high-risk regions in Nicaragua compared to others.^68^ Leibler *et al.* found no association between uKIM-1 or uNGAL with residence in high-risk CKDu regions of Nicaragua.^57^ However, they did identify an association between high uNGAL levels and eGFR <100 ml/min/1.73m^2^.^57^

Additionally, the urine creatinine standardized uKIM-1 and uNGAL levels in Nicaragua were much higher (2 to 7-fold higher) than what was reported for these biomarkers in Sri Lanka despite testing children of similar ages.^57,66,68,78^ For example, the median uKIM-1 for boys was 0.19 ng/mgCr and 0.26 ng/mgCr for girls in endemic areas of Sri Lanka compared to 0.71 ng/mgCr for boys and 1.06 ng/mgCr for girls in endemic regions in Nicaragua.^57,66^ Median uNGAL among girls in Nicaragua was 20.9 ng/mgCr compared to 2.9 ng/mgCr in Sri Lanka, while boys had less variation (4.9 ng/mgCr in Nicaragua and 3.0 ng/mgCr in Sri Lanka). In Mexico, uKIM-1 levels were between those observed in the other studies, but high uNGAL levels were noted in female participants only.^65^

Higher levels of other novel urinary biomarkers (IL-18, MCP-1, YKL-40, cystatin C) were not associated with residence in high-risk CKDu areas.^57,67,68^ Many studies identified differences in novel urinary biomarkers based on sex with females commonly having higher levels than males, although the significance of these differences is unclear.^57,66–68^

Several studies considered the relationship between environmental exposures and kidney injury markers in children. Cárdenas-González *et al.* showed arsenic and chromium exposures are very high in CKDu-endemic areas of Mexico and are associated with higher KIM-1 levels.^65^ Similarly, Nascimento *et al.* showed that children in an agricultural region of Brazil have higher UACR in periods with higher pesticide exposure.^63^ Ismail *et al.* reported that serum creatinine increased immediately following pesticide application both in applicators and those in the community in Egypt.^64^ In Sri Lanka, children with higher heat exposure (based on 5 days/week outdoor sports participation) did not have higher levels of uKIM-1 or UACR.^77^ However, uNGAL was higher in the children with higher heat exposure, but only in the nonendemic CKDu region. In addition, biometeorological data did not indicate a significant difference in thermal indices between CKDu-endemic and nonendemic regions of Sri Lanka.^77^ Notably, the heat index reached levels high enough to require safety precautions for outdoor activities.^77^ This was the only identified study specifically evaluating heat exposure in children.

Renal ultrasounds were evaluated in two studies in Mexico and no urologic anatomic abnormalities were identified.^62,79^ Macias Diaz *et al.* showed that children with persistent albuminuria had lower kidney volume and that those with relatively low kidney volume had decreased eGFR compared to the other participants.^62^ One kidney biopsy was performed by Ortega-Romero *et al.* and revealed Alport syndrome.^79^ Macias Diaz *et al.* reported 18 biopsies, in which glomerulomegaly (72%), partial foot process effacement (100%), microvillous degeneration (80%), and increased organelles (60%) were common findings.^62^ Only one participant had fibrosis and only two participants had a specific histopathologic diagnosis: focal segmental glomerular sclerosis (FSGS) and IgA Nephropathy.^62^

### Kidney Disease Registry and Questionnaire Outcomes

Several analyses of CKD registries revealed the possible burden of CKDu in children. Gutiérrez-Peña *et al.* showed that in Aguascalientes, Mexico, children have a high prevalence of ESKD.^72^ Bukka *et al.* report that the prevalence of kidney disease among primitive tribal groups in India is high overall with the highest prevalence in 6-10 year olds.^75^ Ranasinghe *et al.* showed that CKDu cases of all ages were clustered around paddy fields and irrigation tanks in Sri Lanka.^73^ Two studies in Guatemala by Cerón *et al.* utilized the national pediatric nephrology clinic that cares for most patients with advanced CKD in the nation and found that 43% of participants had CKD with undetermined etiology.^70,71^ While this may suggest a possible burden of CKDu in children, it may be a reflection of limited resources for diagnosis. Participants without a known cause for CKD more commonly lived in municipalities with heavy agricultural practices and their mothers had lower levels of education compared to participants with other forms of CKD, although these findings did not reach statistical significance.^71^

Many studies investigated the association of sociodemographic factors with kidney disease markers (Figure 3). Geographic proximity to agriculture or direct or indirect pesticide exposures was associated with kidney damage markers in 4 studies.^62,64,71,73^ Living in close proximity to a river or irrigation was associated with kidney damage markers in 2 studies^61,73^ ; but had no association in another.^62^ Different drinking water sources were not associated with kidney damage markers in 3 studies.^56,62,79^ Macias Diaz *et al.* reported that breastfeeding and higher body mass index (but within the healthy range) appeared to be protective for CKD.^62^ Male sex, a well-known CKDu risk factors in adults, was not associated with CKD or kidney damage markers in most studies.^56,67,71,79^ Interestingly, the 3 studies that evaluated income level and the 2 that evaluated prematurity found no association of these factors with kidney damage markers.^62,71,79^ Other socioeconomic characteristics were studied and selected results are shown in Figure 3.

**Figure 3:**
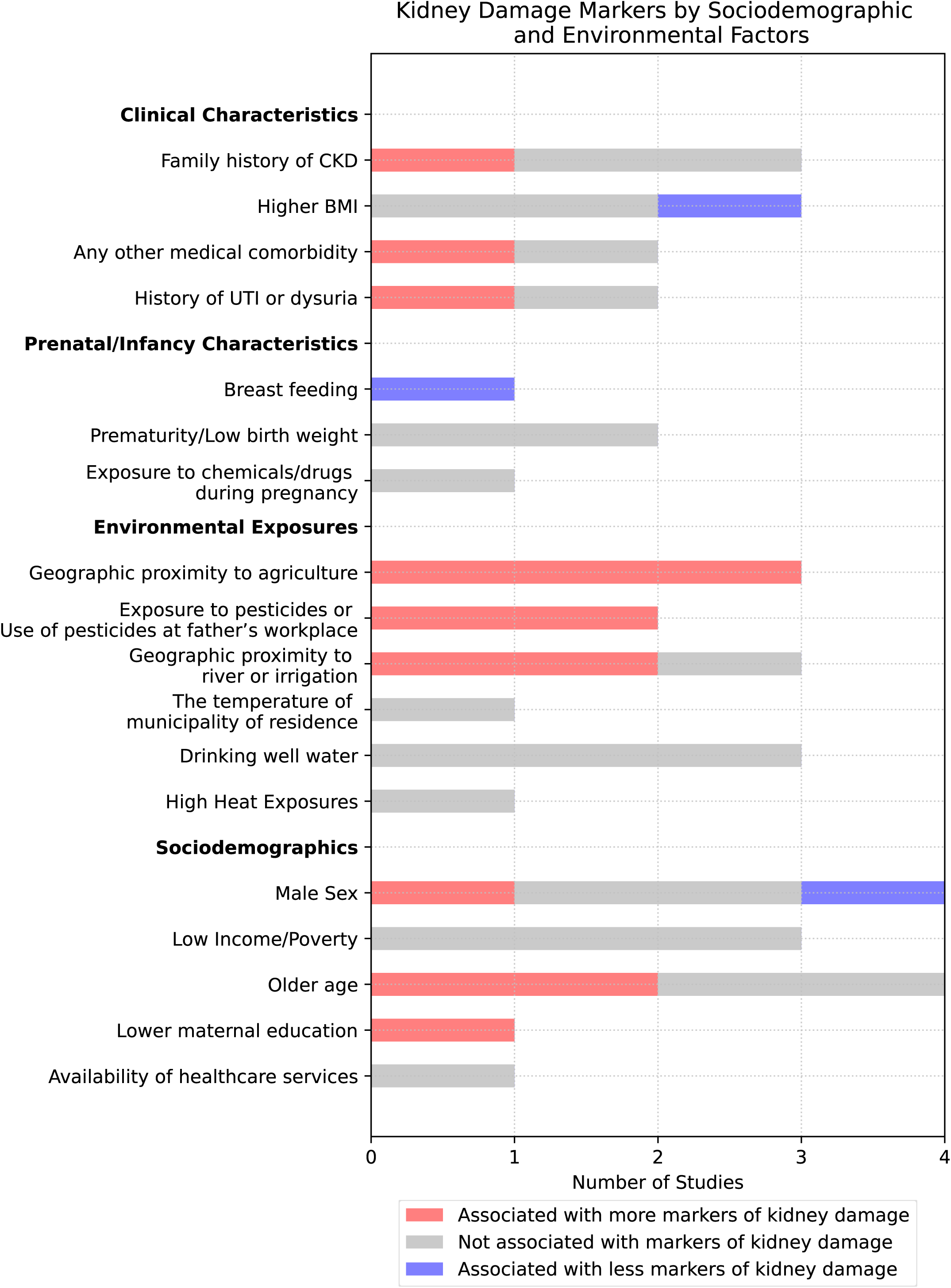
Epidemiologic factors and exposures investigated in included studies. Frequency of studies that investigated selected clinical characteristics, prenatal/infancy characteristics, environmental exposures, and sociodemographics are shown with the association or lack of association of these factors with kidney injury markers.

## Discussion

### Concept of Kidney Injury in Children of Agricultural Communities (KICAC)

While the study designs and results varied, it is remarkable that all 22 studies included across eight countries concluded that there were signs of kidney injury in children living in proximity to areas where CKDu affects adults. Children in CKDu endemic areas demonstrate various signs of early kidney damage, a phenomenon we have termed Kidney Injury in Children of Agricultural Communities (KICAC). KICAC may increase the risk for development of CKDu later in life and may also indicate the presence of the causative agent in the community. The identification of a high prevalence of KICAC in these studies emphasizes the urgent need for ongoing research and preventative efforts even at very young ages and provides critical insight into the potential mechanisms of CKDu.

Despite extensive research on CKDu in adults, our comprehensive review only identified 20 articles and 2 abstracts investigating CKDu in children, even though many CKDu researchers have hypothesized that childhood factors are important in development of disease.^1,12,28,80^ While the included studies all support the existence of KICAC, they exhibit diversity in terms of study population, methodologies, and quality. Consistent findings have not yet been replicated across various populations and regions. Moreover, many kidney disease markers exhibit age, regional, and sex variations, influenced by a range of factors including genetics, nutrition, and the environment.^49,50^

### Interpretation of Included Studies

The included studies had marked limitations (Table 2) as can be expected given the ethical, logistical, and financial challenges of studying children in rural, low-resource settings. Only 7 studies were rated as high quality. Almost all included studies had a cross-sectional design and none of the cohort studies collected biologic samples over more than 3 years. Longitudinal studies are crucial to define the incidence, progression, and effects of disease over time. Another limitation was the lack of following guideline-driven diagnosis of CKD with concurrent serum and urine measurements and confirmation of abnormalities after at least 3 months. Importantly, there are no international guidelines specifically addressing children’s kidney health in these community settings, which is a critical future step. Serum markers of kidney function are less practical in the relatively socioeconomically disadvantaged communities affected by CKDu due to the skills and resources needed and children’s hesitancy about blood sampling. Moreover, serum creatinine is less likely to be abnormal in early stages of kidney injury and it is a less accurate marker in the pediatric population since it is affected by muscle mass.^47^ Further, eGFR equations have not been validated in many of the populations studied and regional/geographic differences influence how well estimating equations approximate measured GFR among pediatric and adult populations.^81,82^

**Table 2:** Strengths and limitations of included studies. Summary of the notable strengths and limitations that contributed to each study’s quality assessment score. Arranged alphabetically.

| Study First Author, year | Strengths | Limitations |
| --- | --- | --- |
| Agampodi<sup>56</sup>, 2018 | -Large sample size in a highly endemic region<br>-Concurrent eGFR measurement for those with albuminuria, compared to a group of age matched controls | -No reference group included for initial urine sampling<br>-Initial urine sample was not a first morning collection. No follow-up sample to confirm persistent albuminuria<br>-Analysis did not define groups with and without albuminuria and compare risk factors |
| Cárdenas-González<sup>65</sup>, 2016 | -Comprehensive analysis of environmental contaminants with concurrent urine and serum markers of kidney dysfunction | -Small sample size<br>-Do not report prevalence data for kidney injury markers<br>-Lack of reference intervals for biomarkers<br>-Single time-point collection of samples<br>-Measurement of toxins in urine could lead to reverse causality<br>-Did not specifically measure the toxic form of contaminants |
| Cerón<sup>70</sup>, 2014 | -Large dataset of patients with a diagnostic work-up for a better classification of CKDu | -Cases could exist undetected or not cared for by this specific clinic, though the clinic is a national referral center<br>-Biopsies were rare, so not all cases may be CKDu by histologic definition<br>-Demographic and environmental exposures not assessed |
| Cerón<sup>71</sup>, 2021 | -Focused on children without definitive etiology of advance CKD with full clinically-indicated work-up<br>-Thorough investigation of demographic data | -Inclusion only of children with very advanced CKD, which is not necessarily representative of the typical case of CKDu<br>-Comparing patients with CKDu to patients with CKD from identified causes limits identification of variables associated with all forms of CKD<br>-Analysis of municipality characteristics is not necessarily representative of each individual participant's exposures |
| De Silva<sup>78</sup>, 2021 | -Large sample size<br>-Identified healthy population for establishing reference intervals<br>-Strong subgroup analysis by age and sex | -Single time point collection of samples<br>-These biomarkers are not validated for clinical use for CKD, and no measurement of eGFR to understand how biomarker values might be associated with eGFR<br>-Regional data not shown |
| García-García<sup>69</sup>, 2020 | -Community-based<br>-Large sample size<br>-Concurrent urine and serum testing<br>-Comparison group of populations in nearby municipalities | -Single time point collection of samples<br>-No environmental exposure data<br>-Lack of analysis of risk factors |
| Gunasekara<sup>6</sup>, 2022 | -Large sample size in established endemic and non-endemic regions<br>-Biomarkers compared to a reference interval established in Sri Lankan population<br>-Clear presentation of results by age, sex, and region | -Single time point collection of urine<br>-No assessment of eGFR |
| Gunasekara<sup>7</sup>, 2023 | -Measurement of multiple biometeorological markers for heat exposure<br>-Comparison of high and low heat exposed groups in both endemic and nonendemic regions | -Single time point collection of urine<br>-No assessment of eGFR<br>-Narrow age range included |
| Gutierrez-Peña<sup>72</sup>, 2021 | -Full registry of patients receiving KRT and biopsy reports in a highly endemic CKDu area<br>-Registry contains information of CKD cause, which is based on clinical work-up | -All histologic results are based on one pathologist's reading<br>-Unclear extent of the clinical work-up for "CKDu"<br>-Only examined patients at single time point<br>-No analysis of risk factors |
| Ismail<sup>64</sup>, 2021 | -Longitudinal, prospective cohort study over 3 years with multiple tests over time<br>-Investigated kidney injury markers in context of timing of pesticide application | -Lack of definition and characteristics of the groups with high and low exposure<br>-Study design changed over time<br>-Lack of demographic and medical information<br>-No eGFR calculation performed<br>-No report of the prevalence of abnormal creatinine<br>-No urine tests |
| Leibler<sup>57</sup>, 2021 | -Tested many urinary biomarkers<br>-Concurrent measurements of urinary biomarkers and eGFR<br>-Included participants from both high-risk and non-endemic areas for CKDu | -Small sample size<br>-No assessment of UACR<br>-Not first void urine sample<br>-Single time point collection<br>-Lack of validated reference intervals for urine biomarkers in this population |
| Lozano-Kasten<sup>60</sup>, 2017 | -Performed 2 first morning assessments of UACR<br>-Concurrent serum analysis in a subgroup<br>-Participants with wide age range and included children from community as well as schools | -30% of the target population did not participate<br>-No comparison/control group<br>-Although this is a follow-up study for a community known to be at high-risk, the participants' results from previous study were not linked.<br>-Did not assess serum tests in entire population<br>-Lack of clinical, demographic, and environmental data |
| Macias Diaz<sup>62</sup>, 2022 | -Included repeat first morning measurements for patients with albuminuria<br>-Concurrent urine and serum tests, kidney ultrasound, and kidney biopsy<br>-Compared ultrasound results to control group<br>-Very thorough questionnaire and clinical work-up<br>-Reported an intervention (Losartan) and outcome | -Pesticide assessment is incomplete and based on parental report |
| Nascimento<sup>63</sup>, 2017 | -Robust environmental toxicity measurement<br>-Measuring at 2 time points representing different levels of pesticide exposures | -Small sample size<br>-No eGFR or urine dipstick measured. Did not report prevalence of albuminuria<br>-Lack of background information on adult CKD in the region |
| Orantes-Navarro<sup>59</sup>, 2016 | -Longitudinal follow-up on abnormalities to validate positive findings after 3 months<br>-Large sample size | -No reference group included<br>-No assessment of clinical or demographic factors or exposures that could be possible risk factors |
| Ortega-Romero<sup>61</sup>, 2019 | -Longitudinal follow-up for 18 months with renal ultrasound and biopsy performed if indicated<br>-Concurrent urine and eGFR collection<br>-Strong and well-described statistical methods | -High loss to follow up<br>-Of the 3 children with indications for biopsy, only 1 accepted |
| Ranasinghe<sup>73</sup>, 2019 | -Large sample size<br>-Recruited patients from the health system and community<br>-Used GPS mapping and determined mortality rates | -Results based on medical record review, which may introduce sampling bias<br>-Most participants had only Stage 1 CKD, which may not carry much clinical significance in older population |
| Ramírez-Rubio<sup>68</sup>, 2016 | -Comparison across multiple regions with different adult CKD burdens<br>-Included UACR and novel urinary biomarkers<br>-Subgroup analysis by age, sex, and regions | -Small sample size<br>-Inconsistent inclusion/exclusion of children whose parents worked in sugar cane across schools<br>-No assessment of eGFR<br>-Single time point urine collection<br>-Collection was not a first morning void, though this is not as much of a limitation for novel urinary biomarkers.<br>-Lack of validated reference intervals for urine biomarkers in this population |
| Sandamini<sup>67</sup>, 2022 | -Large sample size<br>-Selection of healthy children only for establishing reference values<br>-Comparison between clearly defined endemic and non-endemic regions | -No established cutoff values for urinary cystatin C so used UACR as a proxy to characterize abnormal renal function<br>-No assessment of eGFR<br>-Single time-point collection only of urine |
| Strasma<sup>58</sup>,<br>2022 | -Multiple community-based sample from 2 months old to 18 years<br>-Performed urine microscopy | -No reference group included<br>-Single time point collection of samples<br>-Lack of any questionnaire data on exposures or medical history<br>-Lack of description of community differences<br>-No assessment of UACR or of eGFR<br>-Urine microscopy performed by one technician for each sample<br>-Small sample size |

Abbreviations: Estimated glomerular filtration rate (eGFR), chronic kidney disease of unknown etiology (CKDu), chronic kidney disease (CKD), kidney replacement therapy (KRT), urinary albumin to creatinine ratio (UACR), global positioning system (GPS).

Urine assays also face limitations in KICAC. Albuminuria, typically absent in early stages of CKDu, may not be detected in KICAC, and random measurement of total urine protein fails to distinguish kidney disease from benign orthostatic proteinuria.^83^ Many studies lacked an accurate assessment of albuminuria given the difficulties of collecting a first morning urine and repeating the collection after at least 3 months to confirm chronicity.^76^ Other urinary biomarkers seem promising for early detection of tubular injury in children, but their clinical utility and external validity are not yet fully established, even in high-resource settings.^49,50^ Regional variations in urine biomarkers between Sri Lanka and Central America CKDu endemic areas highlights the importance of defining reference intervals for urinary biomarkers in different populations.^67,78^

Longitudinal follow-up and comprehensive diagnostic workups are lacking in many of these studies. Few studies include renal ultrasound or kidney biopsies in children, which could strengthen the hypothesis that KICAC progresses to CKDu. Communities at risk are mostly in low-income regions, so future efforts should prioritize risk-factor mitigation approaches such as the provision of clean water alongside research focused in order to protect kidney health in children living in agricultural areas.

### Markers of Kidney Injury in Children of Agricultural Communities (KICAC)

An important observation from this review is that the kidney injury identified in children in CKDu-endemic areas was usually subclinical under existing paradigms for kidney health assessment. Most study participants were children attending school, and therefore, were presumably not feeling ill. CKD is a silent disease until much later stages, which makes it difficult to diagnose. KICAC was identified in some studies by traditional markers, such as serum creatinine and urine albumin, however, many studies only identified KICAC using urine biomarkers.

Studying novel urinary biomarkers may benefit CKDu research by elucidating mechanisms of disease. For example, uKIM-1 is known to be elevated following toxic insults, such as cadmium exposure, which is also associated with interstitial fibrosis and inflammation.^84,85^ uNGAL is elevated in the setting of tubular dysfunction of many different etiologies including cisplatin-induced tubulopathy and also correlates with interstitial fibrosis and tubular atrophy on biopsy.^86,87^ uNAG is a marker of proximal tubular damage and has been associated with progression of other forms of CKD in children.^88,89^

We did not identify a clear indication for choosing one specific urinary biomarker over the others in the context of KICAC. For biomarkers included in more than one study (uKIM-1, uNGAL, uNAG, uIL-18), the results often differed between studies and regions. Pediatric uKIM-1 levels seemed to be correlated with adult CKDu burden in Sri Lanka^66,77^ and with arsenic and chromium exposure in Mexico^65^ but did not correlate with areas of high adult CKDu burden in Nicaragua.^57^ Urinary NGAL levels were higher in endemic CKDu regions in Nicaragua^57,68^ but not in Sri Lanka.^66^ Urinary NAG levels were related to adult CKDu burden in Nicaragua^68^ but did not correlate with time points of higher pesticide exposure in Brazil.^63^ Urinary IL-18 was not correlated with areas of disease burden in two studies in Nicaragua.^57,68^

The sample collection methods, such as the timing of the urinary samples, were not consistent, which could have influenced these results. Particularly, first morning urine samples are preferred and were not obtained in every study. In addition, variability of biomarker concentration in urine depending on urine output remains a key concern, and adjustment of urinary biomarker levels to creatinine or specific gravity is usually adopted as a corrective measure. As shown by a recent study, absolute concentrations of KIM-1, NGAL, and cystatin C can be adopted for detection of subclinical kidney injury in adolescents in community screening where first morning urine sampling is practiced.^90^

It is notable that many studies demonstrated that girls had higher levels of urinary biomarkers than boys, even though adult males are at higher risk for CKDu than adult females. This could indicate that the presence of certain biomarkers is protective at certain time points or may reflect differential exposure while *in utero,* during childhood, and/or adulthood by sex. For example, it has been shown that KIM-1 expression initially protects against insults, but sustained expression may lead to progressive interstitial inflammation, fibrosis, and worsening kidney function.^85,91^

It is likely that a combined biomarker signature, proteomics, metabolomics, and/or genomics will be more useful for KICAC detection than a single biomarker alone.^49,52,53^ It is also possible that urine biomarkers at a population level may indicate the presence of the causative environmental exposure, even if individual levels are not linked to CKDu. Given that most of these biomarkers are not extensively validated, we recommend that future research combine a combination of novel and traditional (eGFR and albuminuria) markers of kidney disease with longitudinal follow-up. Urine samples provide a more convenient and cost-effective approach in community screening. The development and validation of specific urinary markers to identify KICAC or early stages of CKDu is a crucial step in screening, clinical care, and research.

### Regional Variation

Our literature review revealed some interesting regional phenomena. For example, despite the known endemicity of CKDu in India, there is a paucity of pediatric data related to CKDu with only one identified abstract from India included in this review.^75^ We also identified relevant studies in Brazil and Egypt, which are not traditionally regarded as CKDu endemic areas.^63,64^ It is also noteworthy that the only two studies included from Guatemala included pediatric patients with advanced CKD while most other studies included in this review were conducted among generally “healthy” populations.^70,71^

Studies from Mexico reported a high prevalence of pediatric CKD in certain agricultural areas; however, they had a marked prevalence of albuminuria, which is not a major characteristic of CKDu in other regions. Analysis of a renal biopsy registry in children revealed the most common diagnoses were FSGS and minimal change disease with a very low prevalence of tubulointerstitial nephritis.^72^ This correlates with the disease characteristics seen in adults in that region.^92^ This suggests that some of the factors leading to KICAC and CKDu may differ across regions and lead to different forms of kidney damage.

### Clues to the Pathogenesis of CKDu

Environmental exposures present in the community-at-large are frequently implicated in CKDu, and studying children in these communities provides clues about the earliest impacts of environmental exposures as well as potentially unnoticed risk modifiers. We identified 3 studies that showed children who live in CKDu endemic farming communities had biological markers of agrochemical or toxic metal exposures, and these were correlated with markers of KICAC.^63–65^ Questionnaire data also revealed that geographic proximity to agriculture and pesticides was associated with kidney damage markers.^62,64,71,73^ The total proportion of cardiac output delivered to the kidneys, and thus delivery of certain toxicants to the very metabolically active proximal tubule, increases drastically in early childhood.^31,41^ Importantly, environmental exposures are modifiable and therefore represent critical intervenable risk factors that can be reduced or eliminated with effective policies and risk communication strategies.

Early life exposure to environmental contaminants may increase susceptibility to chronic disease later in life through multiple mechanisms. For example, animal models have demonstrated that not only do toxic metals, such as cadmium, cross the placenta, they also alter signaling pathways at key stages of embryonic development.^93^. In addition, although nephrogenesis ends in the perinatal period and glomerular filtration rate reaches adult values by 2 years of age, the pediatric population is undergoing significant epigenetic maturation, which is dysregulated by environmental stress.^33^ Early life exposure to potential toxicants including metals and agricultural pesticides is associated with genome-wide changes in DNA methylation.^33^ Furthermore, elevated kidney injury biomarkers have been reported in children with high fluoride exposures.^94^ Thus, even chronic low-level exposure during childhood influence kidney health throughout the life course.

Interactive effects of other physiologic and socioeconomic factors may further exacerbate later life CKD risk in children with altered kidney development. For example, both prematurity and low birth weight are risk factors for lifetime risk of CKD, which is thought to be related to low nephron mass and incomplete nephrogenesis.^35,95^ Maternal risk factors for prematurity and low birth weight include rural residence and low family income, which are characteristics of many CKDu endemic areas.^95^ However, there is a paucity of data evaluating these early life risk factors in CKDu endemic areas.^96^ Nephron mass was only assessed indirectly through renal ultrasound by Macias Diaz *et al.* who identified an association with decreased total renal volume and decreased eGFR.^62^ When assessed by questionnaire, prematurity^61,62^ and other pregnancy-related complications^62^ did not show an association with KICAC. Further research is needed to investigate the potential relationship between prenatal exposures, fetal kidney development, and CKDu. The adult CKDu literature often identifies a relationship of disease with occupational exposures such as agrochemicals and heat stress.^1,2^ Notably, despite the association in adults (particularly in Mesoamerica) of CKDu and heat stress, the only study that evaluated pediatric heat exposure showed no association with KICAC in Sri Lanka.^77^

A common perception is that children do not develop kidney disease until working age, however, the studies reviewed here challenge that interpretation (Figure 4). As presented in these studies, we argue that there are deleterious factors such as poor socioeconomic determinants of health, toxic environmental exposures in the community-at-large, increasing exposure to heat stress, and genetic susceptibility to kidney disease that are present throughout the lifespan. Fetal kidney development could be negatively affected by toxic environmental exposures and poor maternal health and nutrition.^36,37,41,97^ Children may develop KICAC that is not detected by traditional markers. Upon entering the workplace in early adulthood, further kidney injuries could occur through occupational toxic exposures, such as agrochemicals, or from heat exposure and volume depletion. In addition, behaviors such as inadequate water intake, intake of sugary beverages, alcohol consumption, smoking, and nonsteroidal anti-inflammatory drug use could also negatively affect kidney health.^98,99^ While some individuals will experience complete kidney recovery after these insults, individuals with a susceptibility to disease from KICAC could be less likely to experience kidney recovery and go on to develop CKDu.

**Figure 4:**
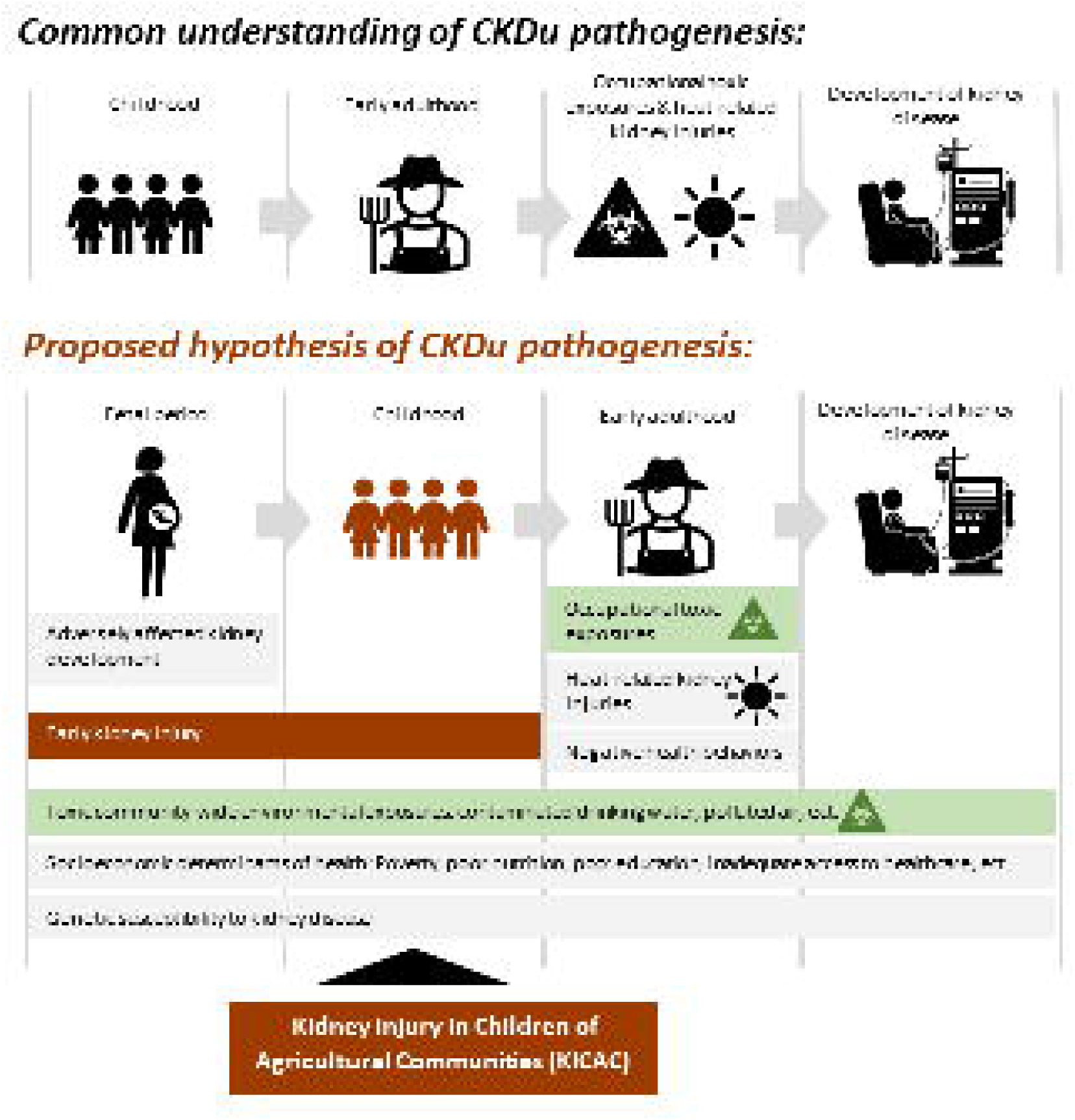
Pathogenesis of CKDu: common understanding versus our proposed hypothesis. CKDu is often attributed to occupational exposures, however, the studies in this review suggest that CKDu pathogenesis may start much earlier and may be detected in the pediatric population through markers of Kidney Injury in Children of Agricultural Communities (KICAC).

### Recommendations

Improving pediatric kidney health in adult CKDu endemic areas is complex and requires involvement of many stakeholders including researchers, communities, and health systems. The studies evaluated here emphasize that children *in utero* and in early childhood should be prioritized in CKDu research. They are a vulnerable population suffering from early kidney damage due to unknown causes that must be identified and addressed. Investigations in the pediatric population appear to have great potential for identifying the etiologic agent(s) of disease. In addition, children may benefit from the screening and follow-up received to reduce later life risk of CKD.

Future analyses should be more robust in methodology and design. Long-term longitudinal cohort studies, including the prenatal period, would be particularly useful. Larger sample sizes are needed, especially in Central America. Sequential concurrent serum and urine measurements, as recommended by KDIGO guidelines for diagnosis of CKD, should be upheld in future studies.^76^ Noninvasive novel urinary biomarkers should be pursued as diagnostic and prognostic tools in the setting of KICAC and CKDu alongside the traditional kidney disease markers such as eGFR and UACR. These urinary biomarker measurements should be standardized with improved methodological reporting of sample collection and assays utilized. For each biomarker, reference intervals for different populations should be established rather than simply comparing to reference intervals. Early-life animal model studies and concurrent biologic and environmental samples may also aid in identification of a toxic environmental exposure.

We advocate for increased screening, improved community resources, and more robust education in CKDu-endemic areas, which are often suffering from poor socioeconomic circumstances and the burden of disease. Pediatric kidney health screening, particularly in communities with a high burden of CKDu among adults, is an important approach to ensure pediatric kidney health through early detection with robust biomarkers. The provision of safe drinking water is essential for all children worldwide for the prevention of multiple illnesses and acts as risk-factor mitigation for CKDu. Moreover, the effects of environmental heat on pediatric kidney health are an area that needs further research, especially in the setting of climate change. Arranging appropriate safety precautions and guidelines for outdoor activities are needed to protect children and adolescents. Strict health and environmental standards placed on industries for appropriate use, storage, and disposal of agrochemicals are necessary to defend vulnerable agricultural communities. Health systems should be strengthened to improve prenatal care, increase kidney disease screening, and increase capacity for kidney disease diagnostics, such as ultrasounds. Finally, community-wide and clinician awareness that children can suffer kidney damage in adult CKDu endemic areas is key for early detection of disease. KICAC and CKDu are community-wide health issues that necessitate multidisciplinary research efforts particularly with longitudinal data to better understand how longstanding exposures impact pediatric kidney health.

### Systematic Review Strengths and Limitations

This is the first systematic review to describe CKDu findings in children. Our approach exhibited several strengths, featuring an exceptionally inclusive search strategy that scrutinized over 1,500 titles and abstracts devoid of geographical or study design restrictions to ensure the identification of all pertinent studies. We also included studies that had any children participants, even if it was largely focused on adults. The screening, data extraction, and quality assessment were conducted by multiple authors employing a validated assessment tool. The limitations of our review include the restriction to articles in English or Spanish only and the heterogenous nomenclature of CKDu. We expanded the search strategy to include all names of the disease we could identify in the literature; however, it is possible that there are other names for this disease or different translations for the names of this disease that we did not include. We made meticulous distinctions between studies addressing CKD of any unknown cause and those explicitly discussing CKDu; nevertheless, there remains the possibility of misinterpreting the understanding of other authors, potentially leading to inadvertent exclusion of relevant studies. Our search criteria depended on authors’ awareness of this emerging disease and explicitly identifying proximity to CKDu cases among the adult population. Studies on general pediatric CKD without mention of CKDu, even in known endemic areas, were omitted. Consequently, it is likely that some studies with valuable insights into CKDu in children were not included. Lastly, the included studies were heterogenous in the reported outcomes precluding direct comparisons of key metrics and performance of statistical analyses.

## Conclusion

We identified 20 peer-reviewed publications and two meeting abstracts that studied the kidney health of children in proximity to areas with a high burden of CKDu among adults. These studies were performed in eight countries and were heterogenous in study design and measured outcomes. Despite their differences, all studies concluded that there were signs of increased burden of CKD or markers of kidney injury in children living in CKDu endemic areas or with a CKDu-relevant exposure. This suggests that the pathophysiologic processes leading to CKDu may begin early in life. Most CKDu literature has concluded that the disease is multifactorial and complex in origin, and the studies in this review strongly suggest that at least some of these factors occur prior to adulthood and possibly even *in utero*. Preventative efforts and research should not focus solely on occupational hazards, but rather focus on factors and exposures that affect entire communities. It is vital to study the pediatric populations in CKDu endemic areas in order to inform strategies for preventative efforts that could mitigate the devastating effects that CKDu has on children, families, and communities.

## Supporting information

Search Strategy

## Data Availability

All studies used this systematic review are publicly available.

## Notes

**<u>Conflicts of Interest:</u>** The authors declare they have no conflicts of interest related to this work to disclose.

### Competing Interest Statement

The authors have declared no competing interest.

### Funding Statement

This study did not receive any funding.

