## Supplementary material for "Pediatric Kidney Injury in Communities Impacted by Chronic Kidney Disease of Unknown Etiology (CKDu): A Comprehensive Systematic Review of Epidemiologic Studies": Search Strategy

**Supplemental Material**

**Search Strategies**

**Librarian Searcher: Elizabeth Blackwood, MSLS; Duke University Medical Center Library & Archives, Duke University School of Medicine**

Peer Review of Search Conducted by: Samantha Kaplan, Ph.D., MLIS, Duke University Medical Center Library & Archives, Duke University School of Medicine

**MEDLINE (via PubMed)
Search date: 1/13/2023**

| Concept | Strategy | Results |
| --- | --- | --- |
| *#1 Chronic Kidney Disease of Unknown etiology (CKDu)* | "Chronic Kidney Diseases of Uncertain Etiology"[Mesh] OR (("Renal Insufficiency, Chronic"[Mesh] OR "chronic kidney disease"[tiab] OR "chronic kidney diseases"[tiab] OR "chronic renal insufficiency"[tiab] OR "chronic renal insufficiencies"[tiab] OR "chronic renal disease"[tiab] OR "chronic renal diseases"[tiab] OR CKD[tiab]) AND (unknown[tiab] OR uncertain[tiab]) AND (etiolog*[tiab] OR aetiolog*[tiab] OR cause[tiab] OR causes[tiab] OR causation[tiab] OR origin[tiab])) OR "Mesoamerican nephropathy"[tiab] OR "Mesoamerican nephropathies"[tiab] OR "CKDnt"[tiab] OR "Chronic interstitial nephritis of agricultural communities"[tiab] OR CINAC[tiab] OR CKDu[tiab] | 2,376 |
| *#2 Agricultural Communities* | ("Renal Insufficiency, Chronic"[Mesh] OR "Chronic Kidney Disease"[tiab] OR "Chronic Kidney Diseases"[tiab] OR "chronic renal insufficiency"[tiab] OR "chronic renal insufficiencies"[tiab] OR "chronic renal disease"[tiab] OR "chronic renal diseases"[tiab] OR "CKD"[tiab]) AND ("Agriculture"[Mesh] OR **agricultur*[tiab] OR farm*[tiab])** | 349 |
| *#3* | #1 OR #2 | 2519 |
| *#4 Pediatrics* | "Hospitals, Pediatric"[Mesh] OR "Child"[Mesh] OR "Infant"[Mesh] OR "Adolescent"[Mesh] OR "Pediatrics"[Mesh] OR "Minors"[Mesh] OR "Puberty"[Mesh] OR infant[tiab] OR infants[tiab] OR infancy[tiab] OR newborn[tiab] OR newborns[tiab] OR neonatal[tiab] OR neonate[tiab] OR neonates[tiab] OR baby[tiab] OR babies[tiab] OR preterm[tiab] OR prematurity[tiab] OR toddler[tiab] OR toddlers[tiab] OR boy[tiab] OR boys[tiab] OR boyhood[tiab] OR girl[tiab] OR girls[tiab] OR girlhood[tiab] OR kid[tiab] OR kids[tiab] OR child[tiab] OR childhood[tiab] OR children[tiab] OR stepchild[tiab] OR stepchildren[tiab] OR schoolchild[tiab] OR schoolgirl[tiab] OR schoolgirls[tiab] OR schoolboy[tiab] OR schoolboys[tiab] OR "school age"[tiab] OR "school aged"[tiab] OR preadolescent[tiab] OR preadolescents[tiab] OR preadolescence[tiab] OR adolescent[tiab] OR adolescents[tiab] OR adolescence[tiab] OR juvenile[tiab] OR juveniles[tiab] OR youth[tiab] OR youths[tiab] OR teen[tiab] OR teens[tiab] OR teenager[tiab] OR teenagers[tiab] OR teenaged[tiab] OR teenage[tiab] OR youngster[tiab] OR youngsters[tiab] OR "young person"[tiab] OR "young persons"[tiab] OR "young people"[tiab] OR puberty[tiab] OR pubescent[tiab] OR pubescence[tiab] OR prepubescent[tiab] OR prepubescence[tiab] OR pediatric[tiab] OR pediatrics[tiab] OR paediatric[tiab] OR paediatrics[tiab] OR minors[tiab] OR PICU[tiab] OR "emerging adult"[tiab] OR "emerging adults"[tiab] | 4,758,178 |
| *#5 Combining* | #3 AND #4 | 428 |
| *Validation String* | **34438575** OR **29769043 OR 32504218 OR 35577796 OR 35155872 OR 34362237 OR 26311057 OR 35107783** | 8/8 |

**CINAHL Complete (via Ebsco)
Search date: 1/13/2023**

| Concept | Strategy | Results |
| --- | --- | --- |
| *#1 Chronic Kidney Disease of Unknown etiology (CKDu)* | MH ("Renal Insufficiency, Chronic" AND ((unknown OR uncertain) AND (etiolog* OR aetiolog* OR cause OR causes OR causation OR origin)) OR TI ("Chronic Kidney Diseases of Uncertain Etiology" OR (("Renal Insufficiency, Chronic" OR "chronic kidney disease" OR "chronic renal insufficiencies" OR "chronic renal disease" OR "chronic renal diseases" OR CKD) AND ((unknownOR uncertain) AND (etiolog* OR aetiolog* OR cause OR causes OR causation OR origin)) OR "Mesoamerican nephropathy" OR "Mesoamerican nephropathies" OR "CKDnt" OR "Chronic interstitial nephritis of agricultural communities" OR CINAC OR CKDu) | 236 |
| *#2 Agricultural Communities* | MH ("Renal Insufficiency, Chronic" AND ((unknown OR uncertain) AND Agriculture OR **agricultur* OR farm*))** OR TI ("Chronic Kidney Diseases of Uncertain Etiology" OR (("Renal Insufficiency, Chronic" OR "chronic kidney disease" OR "chronic renal insufficiencies" OR "chronic renal disease" OR "chronic renal diseases" OR CKD) AND (Agriculture OR **agricultur* OR farm*))** | 159 |
| *#3* | S1 OR S2 | 343 |
| *#4 Pediatrics* | MH ("Hospitals, Pediatric" OR "Child" OR Infant" OR "Adolescent" OR "Pediatrics" OR "Minors (Legal)" OR "Puberty") OR TI (infant OR infants OR infancy OR newborn OR newborns OR neonatal OR neonate OR neonates OR baby OR babies OR preterm OR prematurity OR toddler OR toddlers OR boy OR boys OR boyhood OR girl OR girls OR girlhood OR kid OR kids OR child OR childhood OR children OR stepchild OR stepchildren OR schoolchild OR schoolgirl OR schoolgirls OR schoolboy OR schoolboys OR "school age" OR "school aged" OR preadolescent OR preadolescents OR preadolescence OR adolescent OR adolescents OR adolescence OR juvenile OR juveniles OR youth OR youths OR teen OR teens OR teenager OR teenagers OR teenaged OR teenage OR youngster OR youngsters OR "young person" OR "young persons" OR "young people" OR puberty OR pubescent OR pubescence OR prepubescent OR prepubescence OR pediatric OR pediatrics OR paediatric OR paediatrics OR minors OR PICU OR "emerging adult" OR "emerging adults") | 530,644 |
| *#5 Combining* | S3 AND S4 | 14 |

**Cochrane (via Wiley)
Search date: 1/13/2023**

| Concept | Strategy | Results |
| --- | --- | --- |
| *#1 Chronic Kidney Disease of Unknown etiology (CKDu)* | [mh "Chronic Kidney Diseases of Uncertain Etiology"] OR (([mh "Renal Insufficiency, Chronic"] OR "chronic kidney disease" OR "chronic kidney diseases" OR "chronic renal insufficiency" OR "chronic renal insufficiencies" OR "chronic renal disease" OR "chronic renal diseases" OR CKD) AND (unknown OR uncertain) AND (etiolog* OR aetiolog* OR cause OR causes OR causation OR origin)) OR "Mesoamerican nephropathy" OR "Mesoamerican nephropathies" OR CKDnt OR "Chronic interstitial nephritis of agricultural communities" OR CINAC OR CKDu | 326 |
| *#2 Agricultural Communities* | ([mh "Renal Insufficiency, Chronic"] OR "Chronic Kidney Disease" OR "Chronic Kidney Diseases" OR "chronic renal insufficiency" OR "chronic renal insufficiencies" OR "chronic renal disease" OR "chronic renal diseases" OR CKD) AND (Agriculture OR agricultur* OR farm*) | 187 |
| *#3* | #1 OR #2 | 484 |
| *#4 Pediatrics* | [mh "Hospitals, Pediatric"] OR [mh Child] OR [mh Infant] OR [mh Adolescent] OR [mh Pediatrics] OR [mh Minors] OR [mh Puberty] OR infant OR infants OR infancy OR newborn OR newborns OR neonatal OR neonate OR neonates OR baby OR babies OR preterm OR prematurity OR toddler OR toddlers OR boy OR boys OR boyhood OR girl OR girls OR girlhood OR kid OR kids OR child OR childhood OR children OR stepchild OR stepchildren OR schoolchild OR schoolgirl OR schoolgirls OR schoolboy OR schoolboys OR "school age" OR "school aged" OR preadolescent OR preadolescents OR preadolescence OR adolescent OR adolescents OR adolescence OR juvenile OR juveniles OR youth OR youths OR teen OR teens OR teenager OR teenagers OR teenaged OR teenage OR youngster OR youngsters OR "young person" OR "young persons" OR "young people" OR puberty OR pubescent OR pubescence OR prepubescent OR prepubescence OR pediatric OR pediatrics OR paediatric OR paediatrics OR minors OR PICU OR "emerging adult" OR "emerging adults" | 346,612 |
| *#5 Combining* | #3 AND #4 | 140 |

**Embase (via Elsevier)
Search date: 1/13/2023**

| Concept | Strategy | Results |
| --- | --- | --- |
| *#1 Chronic Kidney Disease of Unknown etiology (CKDu)* | (('chronic kidney failure'/exp OR 'chronic kidney failure' OR "chronic kidney disease" OR "chronic kidney diseases' OR 'chronic renal insufficiency' OR 'chronic renal insufficiencies' OR 'chronic renal disease' OR 'chronic renal diseases' OR 'CKD') AND (('unknown' OR 'uncertain') AND ('etiolog*' OR 'aetiolog*' OR 'cause' OR 'causes' OR 'causation' OR 'origin'))) OR 'Mesoamerican nephropathy' OR 'Mesoamerican nephropathies' OR 'CKDnt' OR 'Chronic interstitial nephritis of agricultural communities' OR 'CINAC' OR 'CKDu' | 9,042 |
| *#2 Agricultural Communities* | ('chronic kidney failure'/exp OR 'chronic kidney failure' OR "chronic kidney disease" OR "chronic kidney diseases' OR 'chronic renal insufficiency' OR 'chronic renal insufficiencies' OR 'chronic renal disease' OR 'chronic renal diseases' OR 'CKD') AND ('agriculture'/exp OR 'agricultur*' OR 'farm*') | 2,821 |
| *#3* | #1 OR #2 | 11,432 |
| *#4 Pediatrics* | ('child'/exp OR 'adolescent'/exp OR 'pediatrics'/exp OR [child]/lim OR [infant]/lim OR [adolescent]/lim OR child:ab,kw,ti OR infant:ab,kw,ti OR infants:ab,ti,kw OR infancy:ab,ti,kw OR infantile:ab,ti,kw OR newborn:ab,ti,kw OR newborns:ab,ti,kw OR neonate:ab,ti,kw OR neonates:ab,ti,kw OR neonatal:ab,ti,kw OR baby:ab,ti,kw OR babies:ab,ti,kw OR preterm:ab,ti,kw OR premature:ab,ti,kw OR prematurity:ab,ti,kw OR todder:ab,ti,kw OR toddlers:ab,ti OR children:ab,ti,kw OR childhood:ab,ti,kw OR kid:ab,ti,kw OR kids:ab,ti,kw OR schoolchildren:ab,ti,kw OR schoolchild:ab,ti,kw OR preadolescent:ab,ti,kw OR adolescent:ab,ti,kw OR adolescence:ab,ti,kw OR youth:ab,ti,kw OR youths:ab,ti,kw OR teen:ab,ti,kw OR teenager:ab,ti,kw OR teens:ab,ti,kw OR teenagers:ab,ti,kw OR teenage:ab,ti,kw OR teenaged:ab,ti,kw OR juvenile:ab,ti,kw OR juveniles:ab,ti,kw) NOT (([young adult]/lim OR [adult]/lim OR [middle aged]/lim OR [aged]/lim OR [very elderly]/lim) NOT ([embryo]/lim OR [fetus]/lim OR [newborn]/lim OR [infant]/lim OR [child]/lim OR [adolescent]/lim)) | 5,098,717 |
| *#5 Combining* | #3 AND #4 | 1,141 |

**Web of Science (via Clarivate)
Search date: 1/13/2023**

| Concept | Strategy | Results |
| --- | --- | --- |
| *#1 Chronic Kidney Disease of Unknown etiology (CKDu)* | TS=((("chronic kidney disease" OR "chronic kidney diseases" OR "chronic renal insufficiency" OR "chronic renal insufficiencies" OR "chronic renal disease" OR "chronic renal diseases" OR CKD) AND (unknown OR uncertain) AND (etiolog* OR aetiolog* OR cause OR causes OR causation OR origin)) OR "Mesoamerican nephropathy" OR "Mesoamerican nephropathies" OR "CKDnt" OR "Chronic interstitial nephritis of agricultural communities" OR CINAC OR CKDu) | 2,016 |
| *#2 Agricultural Communities* | TS=(("Chronic Kidney Disease" OR "Chronic Kidney Diseases" OR "chronic renal insufficiency" OR "chronic renal insufficiencies" OR "chronic renal disease" OR "chronic renal diseases" OR "CKD") AND (**agricultur* OR farm*))** | 389 |
| *#3* | #1 OR #2 | 2,163 |
| *#4 Pediatrics* | TS= (infant OR infants OR infancy OR newborn OR newborns OR neonatal OR neonate OR neonates OR baby OR babies OR preterm OR prematurity OR toddler OR toddlers OR boy OR boys OR boyhood OR girl OR girls OR girlhood OR kid OR kids OR child OR childhood OR children OR stepchild OR stepchildren OR schoolchild OR schoolgirl OR schoolgirls OR schoolboy OR schoolboys OR "school age" OR "school aged" OR preadolescent OR preadolescents OR preadolescence OR adolescent OR adolescents OR adolescence OR juvenile OR juveniles OR youth OR youths OR teen OR teens OR teenager OR teenagers OR teenaged OR teenage OR youngster OR youngsters OR "young person" OR "young persons" OR "young people" OR puberty OR pubescent OR pubescence OR prepubescent OR prepubescence OR pediatric OR pediatrics OR paediatric OR paediatrics OR minors OR PICU OR "emerging adult" OR "emerging adults") | 4,019,894 |
| *#5 Combining* | #3 AND #4 | 210 |

**SciELO (via FAPESP - BIREME)
Search date: 1/12/2023**

| Concept | Strategy | Results |
| --- | --- | --- |
| *#1 Chronic Kidney Disease of Unknown etiology (CKDu)* | (Mesoamerican nephropathy) OR (Mesoamerican nephropathies) OR (Chronic Kidney Diseases of Uncertain Etiology) OR (Chronic Kidney Diseases of Unknown Etiology) OR (Chronic Kidney Diseases of Uncertain Origin) OR (Chronic Kidney Diseases of Unknown Origin) OR (chronic kidney disease AND unknown) OR (chronic kidney diseases AND Unknown) OR (chronic kidney disease AND uncertain) OR (chronic kidney diseases AND uncertain) OR (chronic renal insufficiencies AND unknown) OR (chronic renal insufficiency AND Unknown) OR (chronic renal insufficiencies AND uncertain) OR (chronic renal insufficiency AND uncertain) OR (CKDu) OR (CKDnt) OR (Chronic interstitial nephritis of agricultural communities) OR (CINAC) | 74 |
| *#2 Agricultural Communities* | (Chronic Kidney Disease) OR (Chronic Kidney Diseases) OR (chronic renal insufficiency) OR (chronic renal insufficiencies) OR (chronic renal disease) OR (chronic renal diseases) OR (CKD) AND ((**agricultur*) OR (farm*))** | 42 |
| *#3* | #1 OR #2 | 109 |
| *#4 Pediatrics* | (infant OR infants OR infancy OR newborn OR newborns OR neonatal OR neonate OR neonates OR baby OR babies OR preterm OR prematurity OR toddler OR toddlers OR boy OR boys OR boyhood OR girl OR girls OR girlhood OR kid OR kids OR child OR childhood OR children OR stepchild OR stepchildren OR schoolchild OR schoolgirl OR schoolgirls OR schoolboy OR schoolboys OR "school age" OR "school aged" OR preadolescent OR preadolescents OR preadolescence OR adolescent OR adolescents OR adolescence OR juvenile OR juveniles OR youth OR youths OR teen OR teens OR teenager OR teenagers OR teenaged OR teenage OR youngster OR youngsters OR "young person" OR "young persons" OR "young people" OR puberty OR pubescent OR pubescence OR prepubescent OR prepubescence OR pediatric OR pediatrics OR paediatric OR paediatrics OR minors OR PICU OR "emerging adult" OR "emerging adults") | 104,972 |
| *#5 Combining* | #3 AND #4 | 3 |
